# Evaluating Behavioral Parallelism and Cost Incentives in Voluntary Quarantine: A Randomized App-Based Epidemic Game at VinUniversity in Hanoi, Vietnam

**DOI:** 10.64898/2026.10.06.26364443

**Authors:** Andrés Colubri, Anh Thu Le, Ngoc Le Thi Minh, Vu Nhat Huy Ho, Ngoc Minh Anh Dang, Huong Duong Le, Hoang Minh Doan, Dale King, Jasmina Panovska-Griffiths, Pei Yee Woh

## Abstract

**Background:** Quantifying behavioral drivers of protective action during infectious disease outbreaks remains challenging. App-based experimental epidemic games (epigames) provide a framework for studying these decisions in naturalistic settings, but correspondence between in-game measures and real-life beliefs or behaviors requires evaluation. We examined whether an activity-based opportunity cost influenced voluntary quarantine decisions within an epigame and whether real-life health beliefs transferred into the game and predicted observed choices.

**Methods:** We conducted a two-week randomized controlled trial from May 4 to May 17, 2026, at VinUniversity (VinUni) in Hanoi, Vietnam. Participants using the Epigames app installed in their smartphone were randomized to a low-barrier group, earning 1 point per qualifying contact with other participant when remaining active, or a high-barrier group, earning 2 points per contact. Of 439 enrolled participants, 283 were successfully randomized to one of the two groups, made at least one valid daily quarantine decision, and their data was used in the analysis. Real-life (S1) and in-game (S2) beliefs measured susceptibility, severity, quarantine self-efficacy, and perceived benefits. Belief correspondence was assessed using Holm-adjusted Spearman correlations and prospective cumulative-logit models. Quarantine choices were analyzed using participant-day binomial generalized estimating equations (GEE) and participant-level quasi-binomial models.

**Results:** Participants made 1,823 valid daily decisions and chose quarantine on 458 participant-days (25.1%). Quarantine rates were 30.1% in the low-barrier group and 20.2% in the high-barrier group. The high barrier was associated with 41% lower population-averaged odds of quarantine (GEE odds ratio [OR] 0.59, 95% confidence interval [CI] 0.40–0.86, *p* = 0.0059); the quasibinomial estimate was identical (OR 0.59, 95% CI 0.40–0.86, *p* = 0.0055). Among 251 participants with matched S1–S2 data, all four corresponding beliefs were positively associated (Spearman’s *ρ* = 0.259–0.418; all Holm-adjusted *p* < 0.001). Prospective ordinal models confirmed positive matched-domain associations (ORs 1.39–2.18 per one-point S1 increase). Beliefs showed little robust association with quarantine behavior, and neither S1 nor S2 beliefs jointly modified the randomized effect. Self-efficacy, network degree, and the preregistered gender main effect were also unsupported.

**Conclusion:** A higher activity-based opportunity cost reduced voluntary quarantine within the VinUni epigame, while participants carried relative differences in stated health beliefs from real-life to in-game framing. These beliefs did not reliably predict observed quarantine choices within the game framing. Epigames can support controlled study of incentives and belief correspondence in naturalistic settings such as college campuses, but behavioral validity or real-world generalizability requires further work.

## 1. Introduction

Infectious disease outbreaks are shaped not only by pathogen characteristics and biological transmission processes, but also by human behavior. Individual responses to perceived infection risk such as changes in social contact, physical distancing, and voluntary quarantine, can alter opportunities for disease transmission, thereby influencing epidemic trajectories and the effectiveness of public health interventions [1]. These behavioral responses are dynamic rather than fixed. Individuals may modify their actions as they receive new information, observe changes in disease incidence, perceive different levels of personal risk, or weigh the expected benefits of protective actions against their social and economic costs [2, 3]. However, identifying the causal drivers of protective behaviors remains methodologically challenging. Observational studies provide valuable evidence from real-world outbreaks, but multiple overlapping factors including public health policies, changes in disease incidence, and risk perception often act simultaneously on behavior [4, 5]. These factors, together with the non-random implementation of many interventions, can make it difficult to isolate the effect of a specific behavioral determinant or policy [6].

Digital epidemiology provides new opportunities to address some of these challenges [7–9]. Researchers have increasingly explored such approaches to understand human behavior changes during COVID-19 [10–14]. Serious games and epidemic simulations have been used to explore infectious disease transmission, risk perception, social interaction, and individual decision-making without exposing participants to actual infection [15–18]. Compared with conventional questionnaires or retrospective self-reporting, smartphone-based sensing can capture patterns of proximity and interaction at a temporal and spatial resolution.

App-based epidemic games, or *epigames* [19], offer interactive digital systems to simulate epidemic processes while allowing researchers to observe participant behavior under controlled conditions in field or naturalistic settings [20]. During an epigame, participants can make decisions in response to evolving risks and consequences within an interactive environment, bridging the gap between observational epidemiology, behavioral experimentation, and computational epidemic modeling. However, a key challenge in using epigames as behavioral research tools is determining whether behavior observed in the game reflects real-life behavior – a correspondence known as indicator parallelism [21]. This determination is critical because participants may respond to a simulated pathogen, virtual consequences, or game incentives differently than they would in real life. A strong association between real-life beliefs and corresponding in-game beliefs or behaviors would suggest that the simulated environment captures meaningful behavioral constructs, rather than merely measuring responses to game mechanics. Conversely, weak correspondence may indicate that behavior is substantially influenced by features of the game environment itself.

The health belief model (HBM) provides a useful theoretical framework for examining this relationship [22]. The HBM proposes that health-related behavior is influenced by individuals’ perceptions of susceptibility to a health threat, perceived severity of that threat, perceived benefits of a protective action, and perceived ability to act (self-efficacy), among other factors [23]. These constructs provide a structured way to compare health-related beliefs measured in real-life contexts with corresponding beliefs elicited within a simulated epidemic environment. In an epigame, for example, perceived susceptibility may be represented by perceived risk of virtual infection, while perceived benefits and self-efficacy may be framed around the decision to quarantine. Comparing these constructs across real-life and in-game contexts can therefore provide a basis for evaluating the degree of indicator parallelism between the two settings.

Voluntary quarantine and self-isolation are central non-pharmaceutical measures for reducing infectious-disease transmission, but adoption depends not only on their intrinsic beliefs, but also on whether they have the practical and financial capacity to bear its costs. In a nationally representative UK survey early in the COVID-19 pandemic, reported willingness to self-isolate was high across socioeconomic groups; however, respondents with lower household income and minimal savings were less likely to report being able to self-isolate [24]. This pattern suggests a gap between motivation and behavior: people may endorse isolation while facing income loss, insecure employment, inability to work remotely, or insufficient financial reserves. A subsequent UK scoping review found that perceived value in protecting others facilitated self-isolation, whereas financial hardship and anticipated loss of work or wages reduced adherence and could even discourage COVID-19 testing in the first place [25]. Thus, studying economic barriers is important because it can clarify how material costs alter the translation of prosocial intentions into protective behavior, and can inform interventions that reduce those costs during future outbreaks.

In this study, we aimed to evaluate how dynamic activity-based cost incentives influence voluntary quarantine decisions within a simulated environment. Specifically, we sought to: (1) examine the relationship between HBM constructs measured in real-life and corresponding constructs measured; (2) evaluate whether experimentally manipulated opportunity costs influence the frequency of voluntary quarantine; (3) examine whether participants’ position within the observed social-contact network is associated with their behavioral choices; and (4) assess discrepancies between participants’ stated perceptions and their observed behavior. By combining randomized behavioral incentives, longitudinal digital behavioral measurement, and theoretically informed survey measures, we further evaluate the feasibility of epigames as experimental tools for studying belief-behavior relationships in infectious disease contexts and to inform the development of future epidemic simulation platforms with improved behavioral and technical validity.

## 2. Methods

### 2.1 Overview

The methods of the study builds on previous work conducted at the American University of Iraq-Baghdad (AUIB) in November 2025 [26]. The main motivation of conducting a similar epigame in VinUni was to introduce a dynamic, activity-based reward mechanism designed to make the opportunity cost of remaining active more closely linked to participants’ actual social interactions. The magnitude of this activity-based reward was experimentally varied between randomized groups, creating different opportunity costs associated with the decision to quarantine [27]. This design was intended to make the behavioral trade-off more salient while maintaining an experimentally controlled manipulation of the cost of protective behavior. The VinUni implementation incorporated updated methodology and app feature refinements including participant-controlled avatar customization, structured onboarding and instructions, and the integration of real-world proximity interactions into the game’s reward and transmission mechanics. These features to create a more immersive and enactive environment in which participants can make repeated behavioral decisions while simultaneously experiencing simulated epidemic consequences and opportunity costs through the epigame approach.

### 2.2 Study Design and Participants

We conducted a two-week, single-blind parallel-group randomized controlled trial combined with a longitudinal correlational component at the VinUni campus between May 4 and May 17, 2026. We targeted a sample size of *N* ≥ 150 participants, recruited via campus email networks, digital posters, and student life announcements. Eligible individuals were student volunteers, faculty, or staff members possessing a compatible iOS or Android smartphone. The study protocol was pre-registered on the Open Science Framework (OSF) before the start of data collection.

Figure 1 shows the study’s flow diagram detailing overall and per-arm enrollment. Participants were blinded to their specific group allocation upon digital onboarding. Randomization into Group 1 (Low Barrier) or Group 2 (High Barrier) occurred automatically via an in-app algorithm after user consent. Final incentives were distributed via a post-study lottery for gift cards, in which an individual’s probability of drawing was directly proportional to their final accumulated point total.

**Figure 1.**
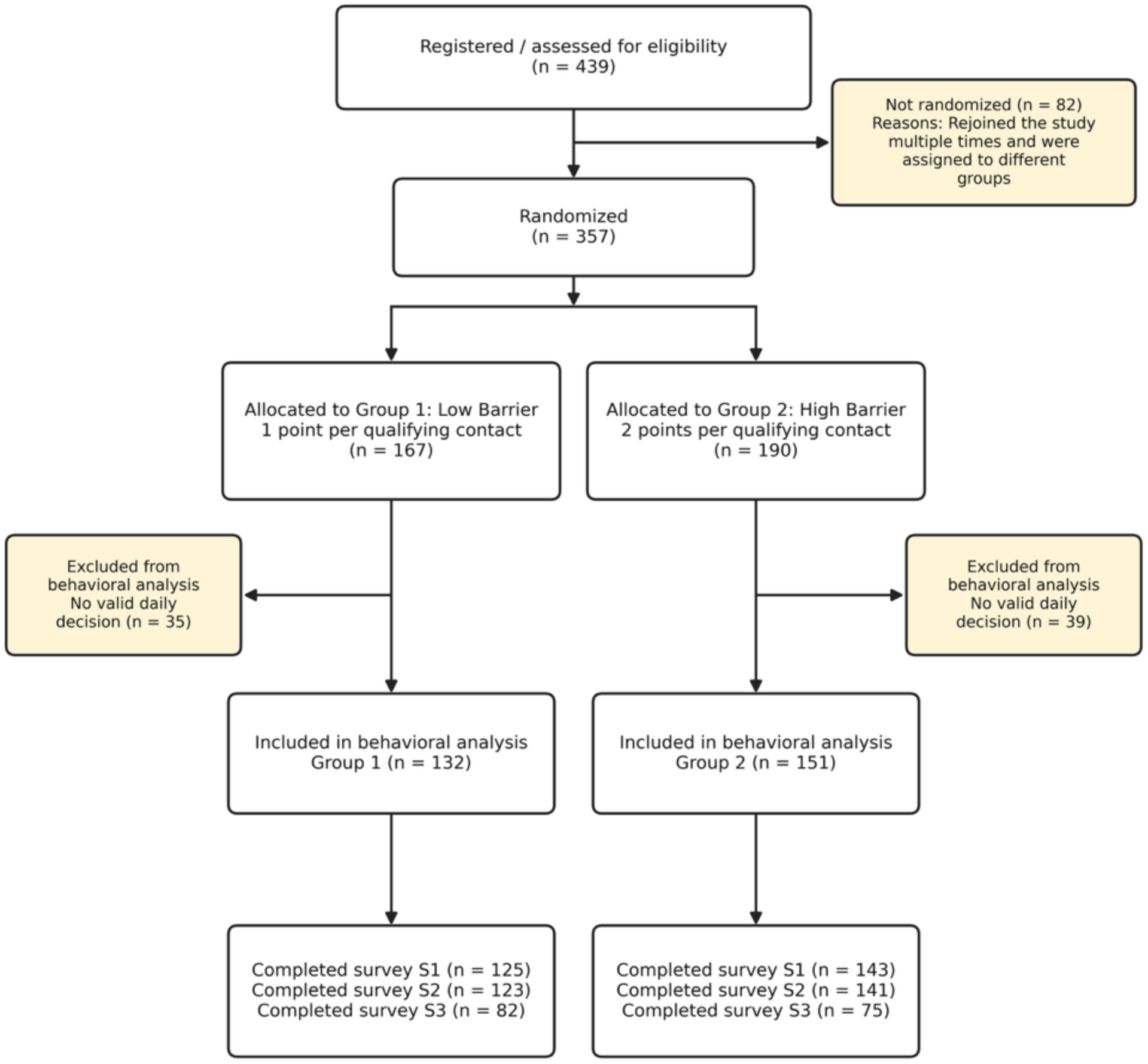
Study’s CONSORT diagram detailing overall and per-arm enrollment of VinUni Epigames.

### 2.3 The Epigames App and Updated Game Mechanics

The study utilized the Epigames mobile application for iOS and Android. This app leverages Bluetooth proximity sensing via the Herald protocol [28] to capture contact networks between participants at high-resolution intervals, simulating the passive transmission of a virtual respiratory pathogen. This iteration introduced several crucial refinements over previous iterations to increase immersion and address prior technical constraints (**Figure 2**):

1. Character Customization: Rather than receiving a pre-assigned avatar, participants utilized a countdown screen prior to launch to review game instructions, followed by a character customization screen where they selected visual features (shape and color) of their digital avatar to foster engagement and user attachment.
2. Dynamic Reward Mechanism: Instead of a static deduction, both arms received a baseline assignment of 5 points daily. Participants choosing to remain active (not quarantining) earned point bonuses for each distinct real-world contact lasting longer than 5 minutes, capped at a maximum of 15 scoring contacts per day.
3. Experimental Manipulation: For Group 1 (Low Barrier), the bonus was set to 1 point per qualified contact. For Group 2 (High Barrier), the bonus was set to 2 points per qualified contact, reflecting the point-gap tension of ∼11 points derived from a Willingness-to-Accept (WTA) pilot conducted at AUIB.
4. Infection Consequences: Transmission resulted in “infection” and “symptoms,” changing the avatar’s visual state and reducing contact point bonuses by a factor of 5 (yielding 0.2 points/ contact in G1 and 0.4 points/contact in G2). Virtual “death” resulted in the total forfeiture of all accumulated points.

**Figure 2.**
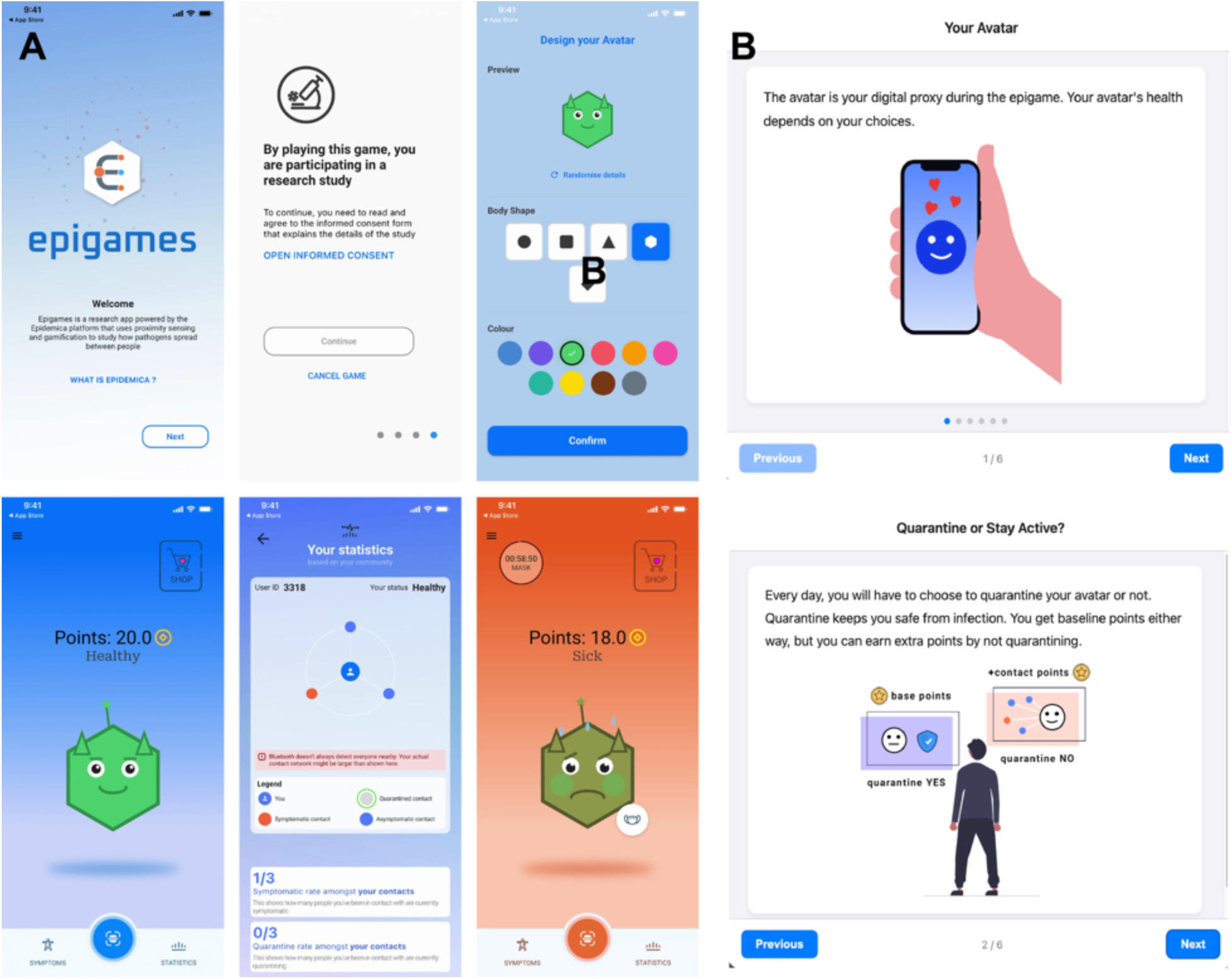
Updated features of the Epigames mobile app used in VinUni trial. (A) Screenshots of the app showing the intro screen, consent form prompt, avatar customization, home screen during the game, statistics page with visualization of player’s immediate network, and home screen with avatar in sick state. (B) Multilingual onboarding tutorial and game instructions, available on the web and through the app.

The study proceeded in structured daily cycles across the 14-day timeline. Each morning, a daily decision window was initiated via a pop-up menu in the application between 12:00 PM and 2:00 PM—a timeframe explicitly selected to minimize conflicts with morning and afternoon university class schedules. Participants selected one of two options:

- Quarantine: Digital proximity scanning was suspended by the app, protecting the participant’s avatar from transmission but restricting their daily yield to the baseline 5 points.
- Do Not Quarantine (Remain Active): Bluetooth scanning remained operational, exposing the avatar to potential infection vectors but enabling the accumulation of contact-based point bonuses (1 point/contact for G1; 2 points/contact for G2).

### 2.4 Survey and Outcome Measures

Three mandatory questionnaires were administered via automated push notifications to capture psychological variables within HBM:

- Survey S1 (Baseline Real-Life Beliefs): Deployed within the first day of the study prior to game interaction. It measured baseline real-world perceptions on a 1–6 Likert scale across four variables: perceived susceptibility to respiratory viruses (S1-Q1), perceived severity of infection (S1-Q2), self-efficacy in executing quarantine (S1-Q3), and perceived effectiveness/benefits of quarantine (S1-Q4), alongside demographic data.
- Survey S2 (In-Game Beliefs): Deployed on or around Day 2 to measure identical HBM constructs reframed specifically around the virtual pathogen environment (e.g., perceived risk of avatar infection and lottery impacts).
- Survey S3 (Exit Survey & Debrief): Administered at the game’s conclusion, utilizing a “cheaptalk” script to mitigate hypothetical bias. It captured perceived game realism (S3-Q1), virtual infection anxiety (S3-Q2), behavioral self-reflection (S3-Q3), and data comprehension (S3-Q4).

The primary outcome was the daily quarantine decision (binary), aggregated per participant into the total number of days quarantined (count) and the overall quarantine rate (proportion of prompts to which the participant responded “quarantine”). One duplicate response by the same participant on the same day collapsed before analysis. Across the analysis population, participants contributed 1,823 distinct participant-days and chose quarantine on 458 of them, an overall quarantine rate of 25.1%. The number of valid decisions per participant ranged from 1 to 12 (mean 6.85 in G1 and 6.09 in G2); this varying denominator was retained explicitly in participant-level models.

Secondary outcomes included correspondence between matched real-life (S1) and in-game (S2) beliefs, modification of the randomized barrier effect by those beliefs, and associations between contact-network degree and quarantine. Daily contacts, realized contact rewards, infection counts, and final point totals were summarized descriptively. The realized daily barrier was defined as the mean contact-point reward available on active participant-days.

### 2.5 Statistical Analysis

Analyses followed the pre-registered hypotheses (all available in the OSF pre-registration page https://osf.io/qev6n/):

- H1 (Main Effect): Participants in Group 1 (Low Barrier) will have a significantly higher overall quarantine rate than “skeptical” participants (those with overall low response scores across all belief variables) in Group 2 (High Barrier).
- H2 (Moderation by cost): The correlation between S1 attitudes (e.g., S1-Q2 Perceived Severity, S1-Q4 Perceived Effectiveness) and quarantine behavior will be significantly stronger in Group 1 (Low barrier) than in Group 2 (High Barrier). We hypothesize the high economic cost in G2 will reduce the influence of personal beliefs.
- H3 (Interaction by efficacy): The difference in quarantine rates between G1 and G2 (the main effect of the barrier) will be moderated by baseline self-efficacy. Specifically, the effect of the higher barrier in G2 will be largest for participants who report low S1 self-efficacy (S1-Q3). Participants with high self-efficacy may be unaffected by cost manipulation, as their behavior is driven by their confidence, not by the barrier.
- H4 (Network effect): Quarantine adoption will be lower among those participants who are hubs in the contact network, as they will experience the highest barrier to adopt the behavior due to its dependence on the number of contacts. This effect will be stronger in G2 than in G1.
- H5 (Gender effect): Quarantine adoption will be higher among participants self-identifying as females.
- H1b, H2b, and H3b, that are verbatim re-statements of H1, H2, and H3 but as function of in-game beliefs S2.

Variance estimation was revised after diagnostic analyses showed substantial extra-binomial variation and within-participant association in repeated daily choices. All tests were two-sided. Estimates are reported with 95% confidence intervals (CIs), and statistical significance was assessed at *α* = 0.05.

Correspondence between real-life and in-game beliefs was assessed using Spearman rank correlations between the four prespecified matched S1–S2 item pairs. Family-wise errors across the four correlations were controlled using the Holm procedure. Because each response was ordinal, prospective cumulative-logit models were also fitted for each S2 item, first using its matched S1 item and randomized group and then using all four S1 items and group. These models were used to assess whether baseline beliefs predicted the corresponding in-game belief and whether other baseline domains added information. Marginal associations between each belief item and participant quarantine rate were summarized with Spearman correlations, group-stratified plots, and Holm-adjusted *p*-values.

The primary participant-level analysis used a binomial generalized linear model with logit link for each participant’s quarantine proportion. The number of valid decisions was supplied as the binomial variance weight, and the model was fitted with Pearson quasi-likelihood scaling (scale=’X2’) to accommodate extra-binomial heterogeneity. This model retained the pre-registered rate-model mean structure and the information contributed by each participant’s number of responses while relaxing the ordinary binomial variance assumption.

In parallel, population-averaged generalized estimating equation (GEE) models were fitted to the participant-day binary decisions, using a binomial family, logit link, participant clustering, exchangeable working correlation, and robust standard errors. GEE represented the disaggregated repeated-decision analysis, whereas the quasi-binomial model represented overall participant quarantine propensity. Agreement between these complementary analyses was used to assess robustness. Unscaled Poisson and binomial models specified in the original analysis plan were not used for final inference because Pearson dispersion was approximately 3 and consecutive choices were strongly associated within participants.

The overall randomized barrier effect was first estimated by models containing group alone. Belief models then included the four S1 items (H1–H3 and H5) or four S2 items (H1b–H3b), binary gender, randomized group, and all belief-by-group interactions. Belief items were mean-centered so that the group coefficient represented the barrier effect on the sample-average belief profile. To evaluate the pre-registered H1 and H1b definitions, separate linear contrasts estimated the group effect for a participant scoring 1 on all four belief items (the specified “skeptic” profile), standardized over the gender distribution of the relevant analysis sample. H2 and H2b were tested with joint Wald tests of the four belief-by-group interactions; H3 and H3b used the corresponding self-efficacy-by-group term. Gender was coded as 0 for men and 1 for women; participants reporting another category or missing gender were excluded only from analyses requiring this binary variable. The pre-registered H5 gender main effect was tested in the multivariable belief models. Group-specific estimates for men and women and the gender-by-group interaction were treated as exploratory analyses. The complete-case S1 and S2 belief models included 251 and 235 participants, respectively.

Hypothesis H4 was evaluated with quarantine propensity modeled as a function of node degree, randomized group, and their interaction. Node degree was each participant’s number of distinct contact partners in the aggregate contact graph. The graph pooled all proximity events and retained an edge when cumulative contact exceeded 0.16 minutes. Degree was calculated before isolated nodes were removed for network-topology analyses so that participants with no qualifying contacts remained in the model at degree zero. Network assortativity described whether participants with similar belief responses were connected more often than expected under bootstrap resampling.

Exploratory transmission-tree analysis used the complete app-attributed transmission graph to preserve infector–infectee chains. The offspring distribution was defined as each infected participant’s number of attributed secondary cases and was fitted with intercept-only Poisson and negative-binomial models. The negative-binomial dispersion parameter was *k* = 1/*α*, where smaller values indicate greater transmission heterogeneity. Following the Lloyd-Smith definition used by Taube et al. [29], a superspreader generated more secondary cases than the 99th percentile of a Poisson distribution with the observed mean [29]. This exploratory graph contained infections excluded from the cleaned daily behavioral series to retain complete chains; its denominator is therefore reported separately and should not be reconciled to the 191 daily infection events in Figure 3.

**Figure 3.**
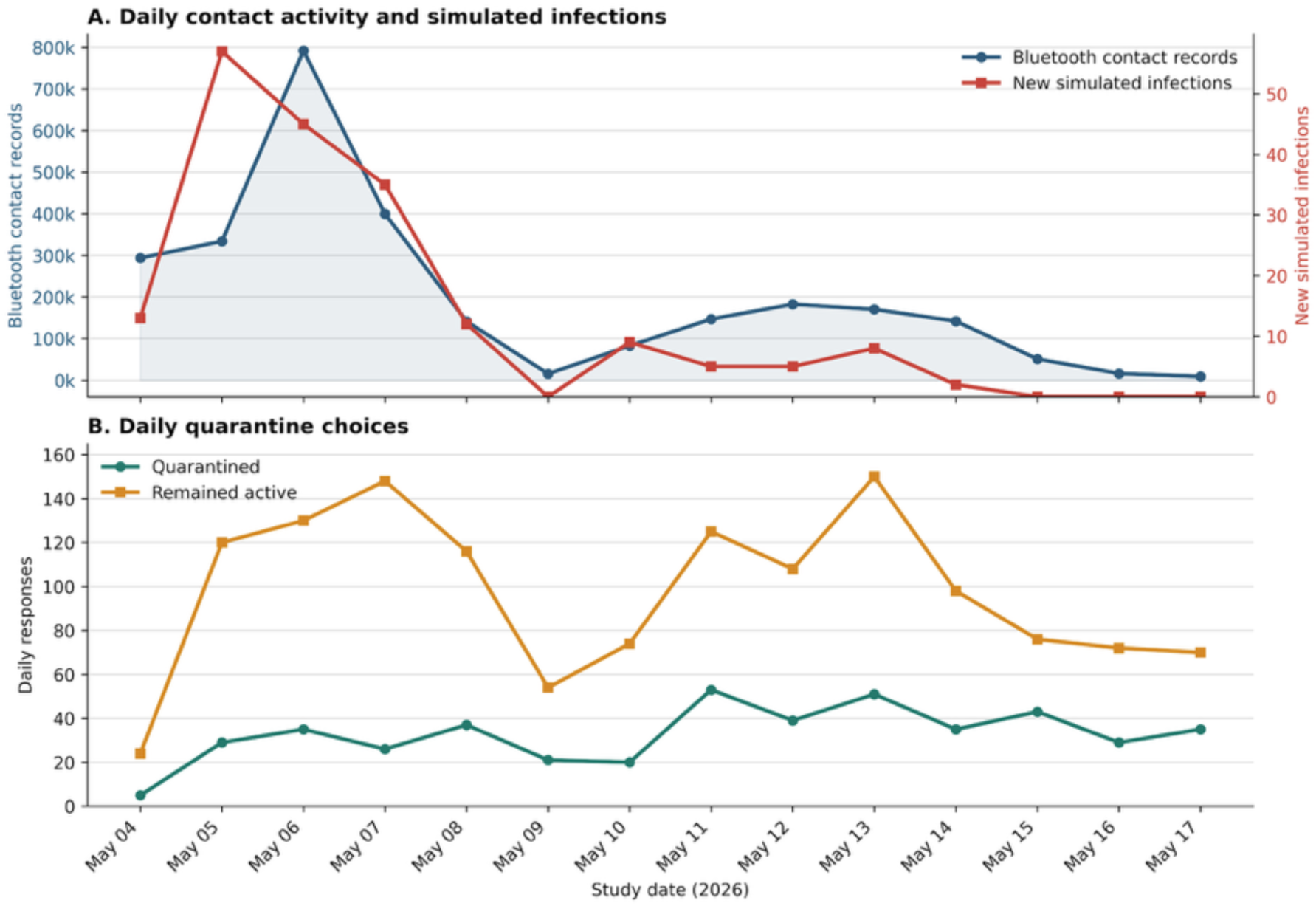
Daily game activity and behavioral choices. (A) Raw Bluetooth contact events and new simulated infection events by study day. The two series use separate vertical scales. (B) Numbers of daily quarantine and remain-active choices. Counts reflect recorded responses and therefore vary with daily participation.

Exploratory analyses examined associations between the four post-game S3 items and quarantine behavior. For each item, participant-level Spearman correlations were supplemented by a participant-day GEE model adjusted for randomized group and a Pearson-scaled participant-level quasi-binomial model with the same adjustment. Separate item-by-group models estimated slopes within G1 and G2 and formally tested heterogeneity between groups. Items were mean-centered within the S3 analysis sample. Holm correction was applied separately across the four items for the Spearman associations, the GEE and quasi-binomial group-adjusted associations, and the GEE and quasi-binomial interaction tests. These post-outcome analyses were not pre-registered and assessed retrospective correspondence rather than prospective prediction.

As sensitivity analyses, the overall group effect was also estimated using fractional logistic regression with HC1 heteroskedasticity-robust errors and ordinary least squares regression of participant quarantine rate with HC3 standard errors. Negative-binomial models were not applied to quarantine proportions because these are bounded binomial outcomes; an exposure-adjusted negative-binomial count model was considered only as a secondary specification check. Analyses used available cases for the variables in each model; model-specific sample sizes are reported.

All analyses were performed in Python 3.13.5 using numpy 2.3.0, pandas 2.3.0, pyarrow 20.0.0, pygraphviz 1.14, matplotlib 3.10.3, networkx 3.5, scikit-learn 1.7.0, scipy 1.15.2, statsmodels 0.14.4, and seaborn 0.13.2. Analysis code is available at https://github.com/colabobio/vinuni-epigame-analysis.

### 2.6 Ethics Approval

The study protocol was approved by UMass Chan Institutional Review Board (study ID 2465) and the VinUni-VinMec Institutional Ethical Review Board for Biomedical Research (approval no. 250/2025/CN/HDDD VMEC). All participants provided digital consent before joining the study through the Epigames app. The study was conducted in accordance with the Declaration of Helsinki.

## 3. Results

### 3.1 Daily Contact, Infection, and Quarantine Patterns

Daily Bluetooth contact activity and simulated infections varied substantially over the 14-day study (**Figure 3A**). The app recorded 2,777,724 raw Bluetooth contact events and 191 new simulated infection events. Contact activity increased from 293,950 events on May 4 to a maximum of 792,084 on May 6, then declined sharply to 15,611 on May 9. New infections peaked one day earlier, at 57 on May 5, remained elevated through May 8, and fell to zero on May 9. A smaller mid-study increases in contact activity occurred from May 10 to May 14, accompanied by 2–9 new infections per day, after which both series tapered; no new infections were recorded during the final three days. These contact counts represent raw proximity records rather than distinct contact partners or contacts meeting the five-minute reward threshold.

Transmission was strongly heterogeneous in the complete app-attributed graph (**Supplementary Table S7; Supplementary Figures S2–S3**). Across three transmission trees, 206 of 288 infected participants (71.5%) had no attributed secondary cases, whereas 36 participants (12.5%) accounted for 228 of 285 person-to-person transmissions (80.0%). A negative-binomial offspring model estimated *k* = 0.176, close to the median *k* = 0.14 reported for COVID-19 transmission trees in the OutbreakTrees database [29]. The mean observed secondary case count was 0.99; because a completed rooted tree with *n* nodes necessarily has *n* − 1 edges, this quantity is structurally constrained toward 1 and was not interpreted as biological *R*_0_.

Daily quarantine-choice counts also fluctuated across the study (**Figure 3B**), partly reflecting variation in the number of participants responding to the daily prompt. Total daily responses ranged from 29 on May 4 to 201 on May 13. Participants selected the quarantine option 5–53 times per day and the remain-active option 24–150 times per day; the remain-active option was more common on every study day. The proportion of daily responses selecting quarantine ranged from 14.9% on May 7 to 36.1% on May 15. Quarantine proportions were generally higher during the second half of the study, even as contact activity and simulated infections declined. These unadjusted temporal patterns describe the aggregate game trajectory and do not by themselves identify the effects of randomized group, infection experience, or study day on individual decisions.

### 3.2 Overall Survey Response Patterns

Figure 4 illustrated complete responses to all four items were available for 268 participants for the baseline real-life belief survey (S1), 264 for the in-game belief survey (S2), and 157 for the exit survey (S3). At baseline, perceived susceptibility was concentrated toward the lower end of the six-point scale (53.4% selected 1 or 2), while perceived severity was concentrated in the middle (59.7% selected 3 or 4). Self-efficacy and perceived benefits were more strongly concentrated at the upper end (52.2% and 67.2%, respectively, selected 5 or 6). The corresponding S2 responses were less polarized: susceptibility and severity remained predominantly mid-scale, while self-efficacy and perceived benefits retained a shift toward higher responses.

**Figure 4.**
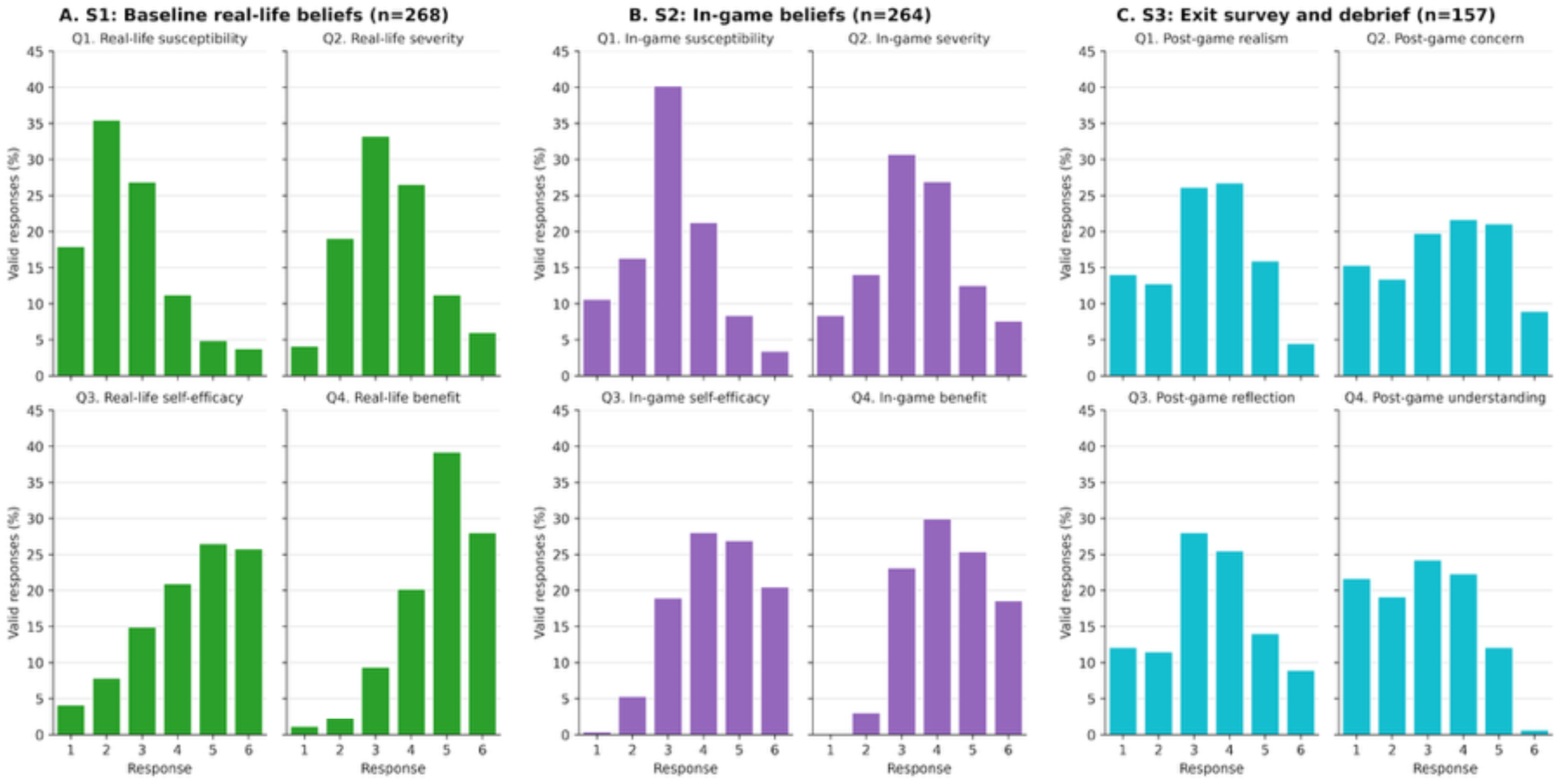
Distributions of valid responses to Questions 1–4 from (A) the baseline real-life belief survey (S1), (B) the in-game belief survey (S2), and (C) the exit survey and debrief (S3). Bars show percentages within each item on the six-point response scale. S1 and S2 assess susceptibility, severity, self-efficacy, and perceived benefits; S3 assesses game realism, concern about virtual infection, reflection on quarantine decisions, and difficulty understanding the information. Panel sample sizes indicate participants with complete responses to all four displayed items.

Exit-survey ratings of game realism, concern about virtual infection, and reflection on quarantine decisions were broadly distributed, with their largest combined shares in responses 3 and 4. Ratings of difficulty understanding the information were shifted lower: 40.8% selected 1 or 2, whereas 12.7% selected 5 or 6. S3 completion was substantially lower than S1 and S2 completion, and these unadjusted distributions are descriptive; they do not establish within-participant changes or between-survey differences.

### 3.3 Participants and Quarantine Decisions

The behavioral analysis included 283 participants who made at least one valid daily decision: 132 in G1 and 151 in G2. The available export did not permit the remaining attrition to be separated into withdrawals and participants who installed the application but never responded to a daily prompt (**Figure 1**). Complete S1 belief data were available for 268 participants, complete S2 data for 264, and matched complete S1–S2 data for 251.

Participants chose quarantine on 458 of 1,823 valid participant-days (25.1%). G1 participants quarantined 272 of 904 participant-days (30.1%), compared with 186 of 919 participant-days (20.2%) in G2 (**Figure 5A**). The GEE estimated 41% lower population-averaged odds of quarantine in G2 than G1 (odds ratio [OR] 0.59, 95% CI 0.40–0.86, *p* = 0.0059). The participant-level quasibinomial estimate was nearly identical (OR 0.59, 95% CI 0.40–0.86, *p* = 0.0055). On the absolute scale, an OLS-HC3 sensitivity model estimated a 9.9-percentage-point lower quarantine rate in G2 (95% CI 2.5–17.2 percentage points lower, *p* = 0.0090).

**Figure 5.**
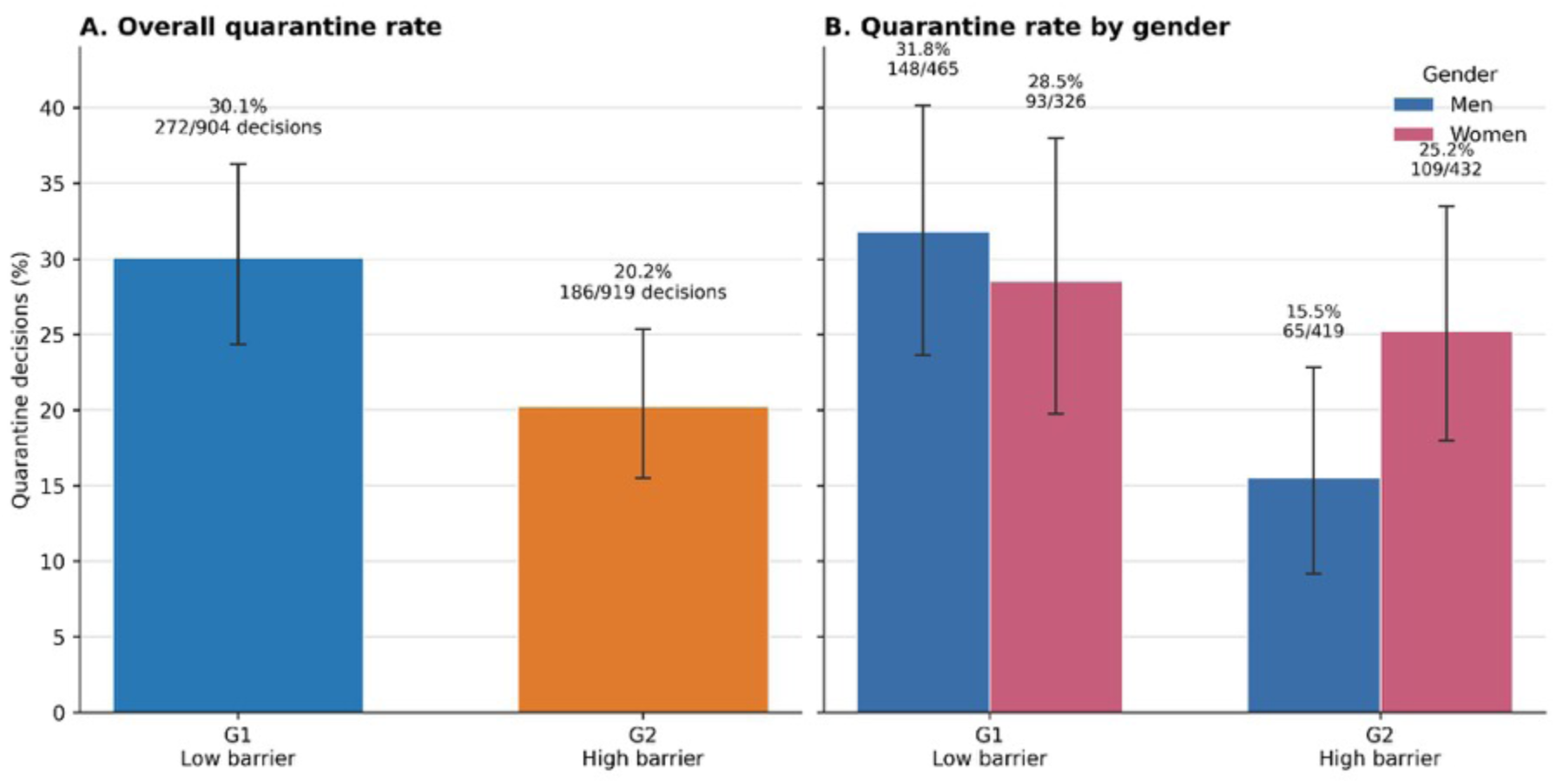
Observed quarantine-decision rates by randomized group. (A) Overall rates in G1 (low barrier) and G2 (high barrier). (B) Rates among men and women within each randomized group. Bars show the ratio of total quarantine decisions to total valid daily decisions; labels give the percentage and numerator/denominator. Error bars are 95% participant-cluster bootstrap confidence intervals obtained by resampling participants within each displayed stratum, thereby retaining each participant’s repeated decisions together. Bars and bootstrap intervals are descriptive; the GEE and quasi-binomial models provide inferential estimates. The gender-stratified pattern was exploratory, and the gender-by-group interaction was borderline rather than conclusive (*p* =0.058).

The ordinary variance assumptions did not describe the data adequately. Pearson dispersion was approximately 3.0–3.5 across the pre-registered binomial and Poisson specifications. Among 1,067 consecutive-decision pairs, the probability of quarantine was 62.6% following a quarantine decision and 14.1% following an active decision (odds ratio 10.23, *p* < 0.001). Exchangeable working correlations in the GEE models ranged from 0.37 to 0.42. These diagnostics supported inference based on the quasi-binomial and robust GEE standard errors. Detailed overdispersion, boundary-mass, and consecutive-decision diagnostics are reported in **Supplementary Tables S4– S5** and **Supplementary Figure S1**.

### 3.4 Realized Reward Manipulation

The implemented reward per qualifying contact closely followed the randomized schedule: the median was 0.959 points in G1 and 1.973 points in G2, a ratio of 2.06 (*p* < 0.001) (**Supplementary Table S7**). The number of qualifying contacts per active day did not differ detectably between groups (1.458 in G1 and 1.333 in G2, *p* = 0.447). Consequently, the realized mean contact reward available per active day was 1.141 points in G1 and 2.325 points in G2, an absolute difference of 1.184 points per day and a ratio of 2.04 (*p* = 0.079). Thus, the realized absolute difference was substantially smaller than the approximately 11-point behavioral-tension target used to motivate the design, although the intended twofold relative reward contrast was implemented. Final point totals did not differ detectably between groups (median 46.70 in G1 and 47.57 in G2, *p* = 0.414).

### 3.5 Transfer of Real-Life Beliefs into the Game

All four matched real-life and in-game belief measures were positively associated among the 251 participants with complete S1–S2 data (**Figure 6**). Spearman correlations were *ρ* = 0.418 for susceptibility, *ρ* = 0.330 for severity, *ρ* = 0.271 for self-efficacy, and *ρ* = 0.259 for perceived benefits; all remained significant after Holm correction (*p_Holm_* < 0.001). Prospective cumulative-logit models gave corresponding odds ratios of 2.18 (95% CI 1.77–2.69), 1.83 (95% CI 1.48–2.26), 1.39 (95% CI 1.18–1.65), and 1.51 (95% CI 1.21–1.88) per one-point increase in the matched S1 item. In each model the matched S1 association remained positive after adjustment for the other three S1 domains and randomized groups. Adding the other baseline domains did not improve prediction of any S2 outcome after Holm correction. These results indicate consistent construct-specific belief transfer from real-life framing to the in-game framing.

**Figure 6.**
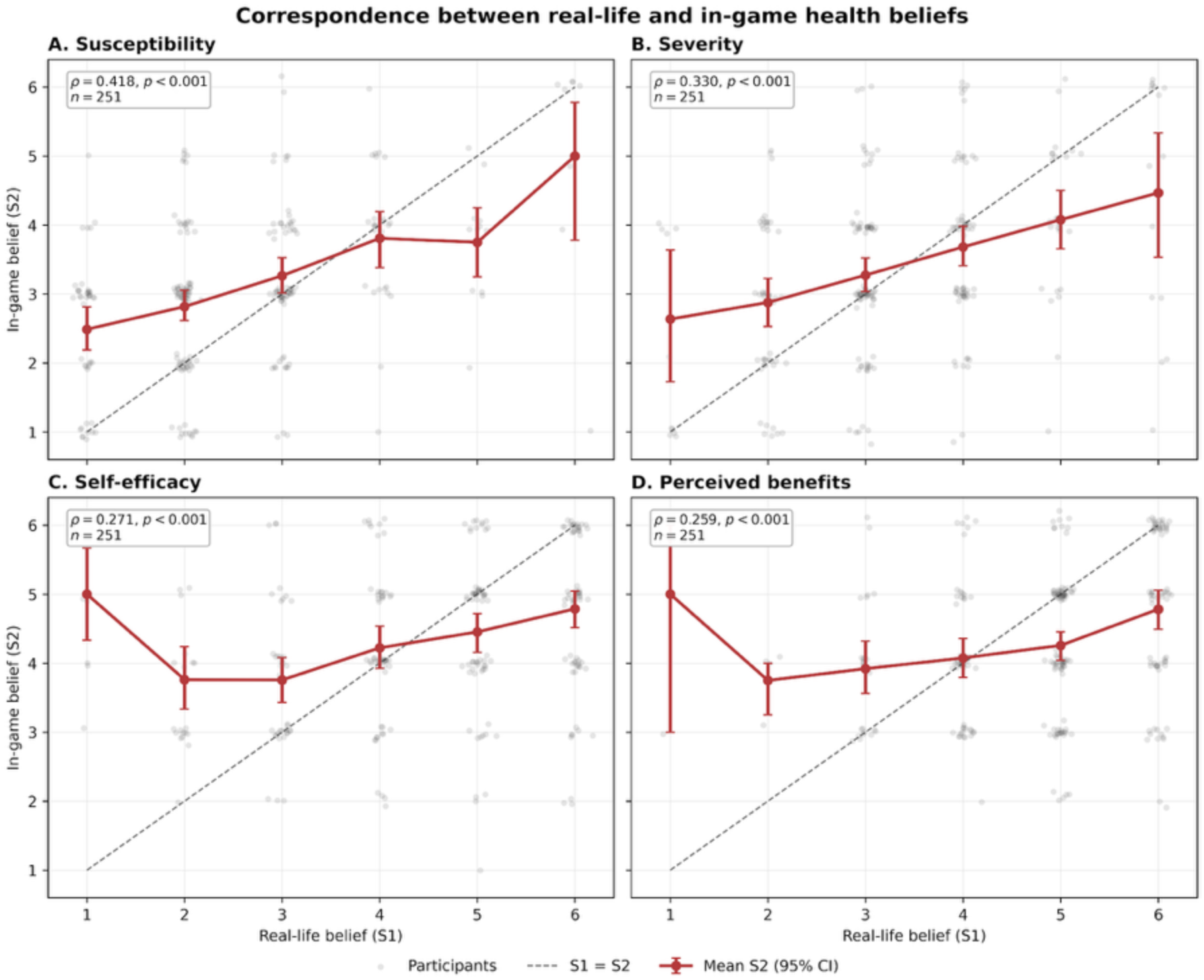
Correspondence between matched real-life (S1) and in-game (S2) health beliefs for (A) susceptibility, (B) severity, (C) self-efficacy, and (D) perceived benefits. Gray points are individual paired responses; jitter was added only for visualization. Red points and lines show the mean S2 response at each S1 level with 95% bootstrap confidence intervals. The dashed line denotes identical S1 and S2 responses. Insets report Spearman rank correlations and paired sample sizes; all four associations remained significant after Holm correction (*p_Holm_* < 0.001).

### 3.6 Beliefs and Quarantine Behavior

In contrast to the consistent S1–S2 correspondence, belief measures showed little association with observed quarantine behavior. In unadjusted participant-level comparisons, S1 perceived severity showed a weak positive trend with quarantine rate (*ρ* = 0.117, unadjusted *p* = 0.056, Holm-adjusted *p* = 0.224) (**Figure 7**). S1 susceptibility, self-efficacy, and benefits were not associated with quarantine rate (all unadjusted *p* ≥ 0.264). In the adjusted GEE model, the S1 severity coefficient was also borderline (log-odds coefficient 0.171, 95% CI −0.005–0.347, *p* = 0.057), whereas the quasi-binomial estimate was smaller and nonsignificant (*p* = 0.172). This pattern was therefore treated as suggestive rather than confirmatory.

**Figure 7.**
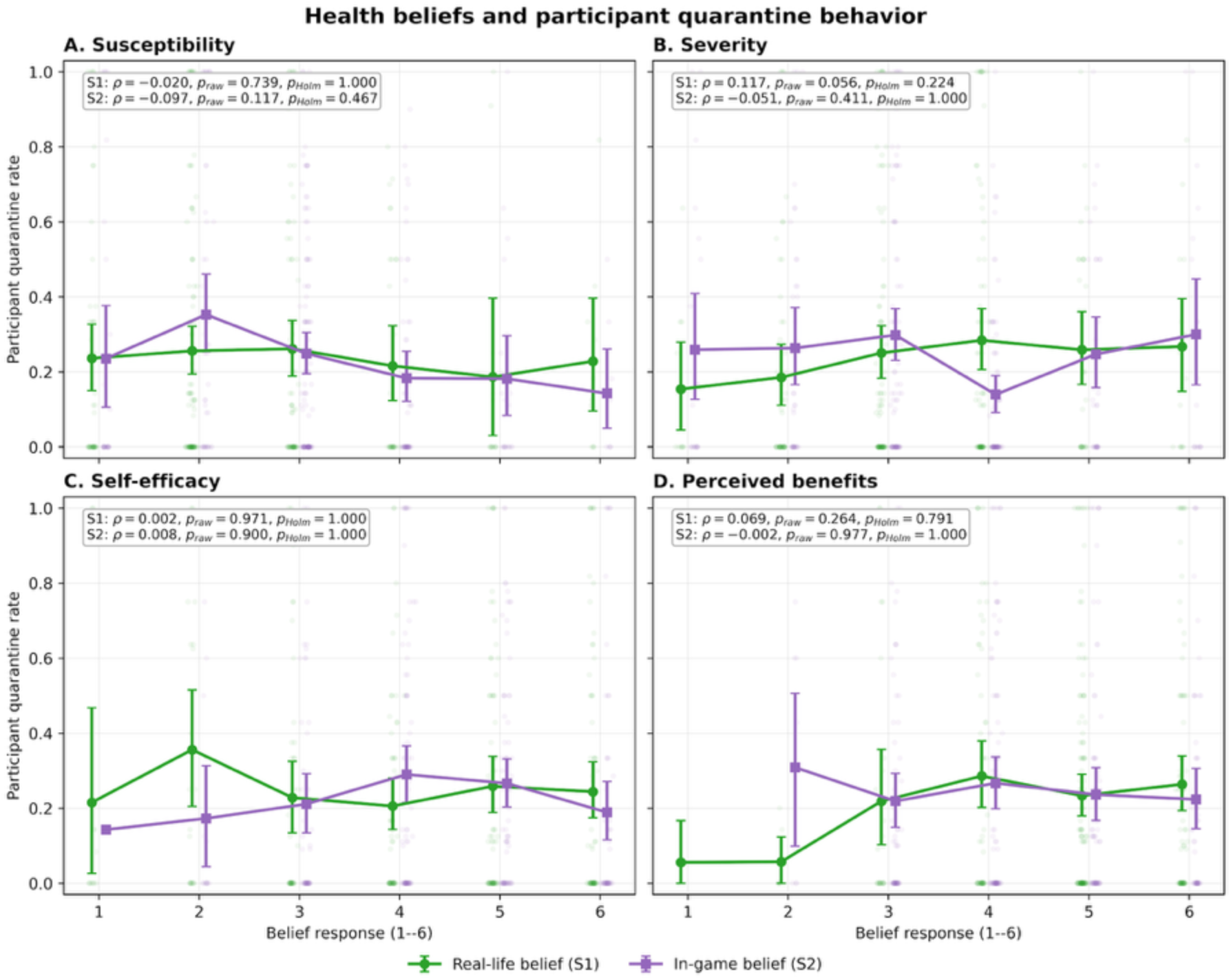
Unadjusted participant-level relationships between quarantine rate and (A) susceptibility, (B) severity, (C) self-efficacy, and (D) perceived benefits. Blue circles represent real-life beliefs (S1), and red squares represent in-game beliefs (S2). Points show individual participants with small horizontal offsets and jitter added only for visualization; lines show the mean quarantine rate at each belief response with 95% bootstrap confidence intervals. Insets report Spearman rank correlations and Holm-adjusted *p*-values, corrected across the four domains separately for S1 and S2. The plots pool randomized groups and are descriptive; adjusted belief and belief-by-group effects were estimated using the prespecified GEE and quasi-binomial models.

The pre-registered belief-dependent barrier hypotheses were not supported under either final specification. For the all-low S1 “skeptic” profile, the standardized GEE barrier contrast was OR 0.50 (*p* = 0.378), with a quasi-binomial OR of 0.57 (*p* = 0.509); H1 was not supported. The corresponding S2 contrast was also null (GEE OR 1.72, *p* = 0.545; quasi-binomial OR 1.62, *p* = 0.599), providing no support for H1b. The four belief-by-group interactions were jointly null for both S1 (GEE Wald *χ*^2^_4_ = 1.10, *p* = 0.895; quasi-binomial *p* = 0.826) and S2 (GEE Wald *χ*^2^_4_ = 2.82, *p* = 0.589; quasi-binomial *p* = 0.351), providing no support for H2 or H2b. Self-efficacy did not modify the barrier effect for either S1 (GEE *p* = 0.382; quasi-binomial *p* = 0.329) or S2 (GEE *p* = 0.292; quasi-binomial *p* = 0.505), providing no support for H3 or H3b. Complete hypothesis tests and individual interaction coefficients are reported in **Supplementary Tables S2–S3**.

### 3.7 Gender-Stratified Barrier Estimates

The pre-registered H5 main effect of gender was not supported in either the S1 model (GEE *p* = 0.295; quasi-binomial *p* = 0.437) or the S2 model (GEE *p* = 0.353; quasi-binomial *p* = 0.412). Exploratory stratified analyses nevertheless showed different point estimates by gender. Among men, the participant-day quarantine rate was 31.8% in G1 and 15.5% in G2; the GEE barrier estimate was OR 0.37 (95% CI 0.21–0.66, *p* = 0.00069). Among women, the corresponding rates were 28.5% and 25.2% (**Figure 5B**), and the GEE estimate was OR 0.81 (95% CI 0.46–1.44, *p* = 0.479). Quasi-binomial estimates were similar for men (OR 0.39, 95% CI 0.22–0.70, *p* = 0.0015) and women (OR 0.85, 95% CI 0.48–1.48, *p* = 0.555). The formal gender-by-group interaction was borderline rather than conclusive (GEE OR 2.19, 95% CI 0.97–4.93, *p* = 0.058). The stratified pattern is therefore reported as exploratory and does not establish that women were unaffected by the barrier. The pre-registered gender main-effect estimates are included in **Supplementary Table S2**.

### 3.8 Contact-Network Position

Neither node degree nor its interaction with randomized group predicted quarantine propensity. In the GEE model, the degree main effect had *p* = 0.243 and the degree-by-group interaction had a log-odds coefficient of 0.005 (95% CI −0.005–0.016, *p* = 0.338); quasi-binomial results were similar (interaction *p* = 0.315). H4 was therefore not supported. Full GEE and quasi-binomial degree-by-group results are reported in **Supplementary Table S2**.

Among the 157 participants with complete S3 data, the mean exploratory dissonance score was 0.213 in G1 and 0.246 in G2. The distributions did not differ in the prespecified direction (Mann– Whitney *p* = 0.747).

### 3.9 Post-Game Reflection and Quarantine Behavior

In exploratory analyses of the 157 S3 respondents (1,238 participant-days), higher S3-Q3 ratings of correspondence between game quarantine decisions and intended real-life behavior were associated with more observed quarantine. The pooled participant-level association was *ρ* = 0.283 (Holm-adjusted *p* = 0.0013). Adjusting for randomized group, the GEE estimated an OR of 1.41 per one-point increase in S3-Q3 (95% CI 1.16–1.71; Holm-adjusted *p* = 0.0019); the quasibinomial estimate was similar (OR 1.39, 95% CI 1.16–1.67; Holm-adjusted *p* = 0.0013).

The association differed by randomized group (**Figure 8**). The GEE slope was OR 1.13 per point in G1 (95% CI 0.88–1.45, *p* = 0.334) and OR 1.88 in G2 (95% CI 1.41–2.50, *p* < 0.001). The ratio of these slopes was 1.66 (95% CI 1.14–2.42; interaction Holm-adjusted *p* = 0.0338). The quasibinomial interaction was similar (ratio of ORs 1.61, 95% CI 1.11–2.33; Holm-adjusted *p* = 0.0469). Associations for perceived realism (S3-Q1), concern about virtual infection (S3-Q2), and difficulty understanding game information (S3-Q4) were not consistently significant after correction across items and model families (**Supplementary Table S1**).

**Figure 8.**
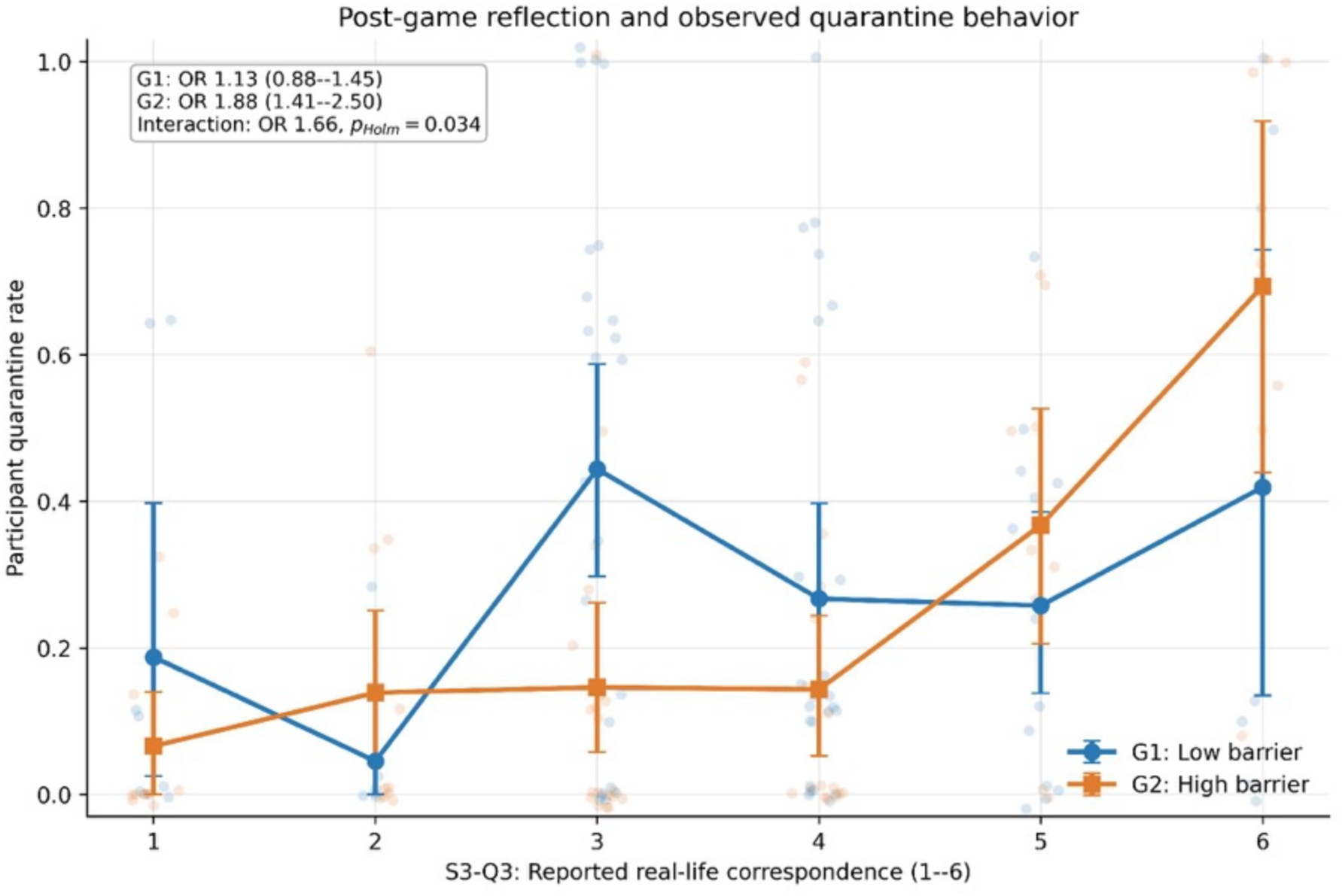
Exploratory association between S3-Q3 retrospective real-life correspondence ratings and participant quarantine rate, stratified by randomized group. Transparent points show participants with jitter added only for visualization; lines and symbols show the mean participant quarantine rate at each response level with 95% participant-bootstrap confidence intervals. Insets report arm-specific participant-day GEE odds ratios per one-point increase and the Holm-adjusted item-by-group interaction. S3 was administered after gameplay, so the association may reflect retrospective consistency, recall, or selection and should not be interpreted as a causal or prospective effect.

S3 completion was selective. Complete responses were available for 82 of 132 G1 participants (62.1%) and 75 of 151 G2 participants (49.7%; *χ*^2^_1_ = 4.42, *p* = 0.0355). Completers also contributed more valid daily decisions than non-completers (means 7.89 versus 4.64; Mann– Whitney *p* < 0.001). The S3 associations therefore describe a more engaged subset and may be affected by post-randomization selection.

### 3.10 Summary of Hypothesis Tests

The randomized high-barrier condition reduced quarantine overall, but the pre-registered prediction that this effect would be concentrated among participants with uniformly low beliefs was not replicated. H1/H1b, the joint belief-moderation tests H2/H2b, the self-efficacy interactions H3/ H3b, the network moderation hypothesis H4, and the H5 gender main effect were all null under the GEE and quasi-binomial specifications. The data instead showed a marked contrast between consistent transfer of matched beliefs from S1 to S2 and little evidence that those beliefs predicted observed quarantine choices.

## 4. Discussion

### 4.1 Principal Findings

This randomized epigame study produced a clear contrast between the transfer of stated beliefs and the prediction of observed behavior. Participants’ baseline real-life beliefs were consistently associated with the corresponding beliefs reported after entering the simulated epidemic environment. This correspondence was domain-specific. Each S1 construct predicted its matched S2 construct, and including the other baseline belief domains did not materially improve the S2 models. Yet neither the real-life nor in-game belief measures reliably predicted participants’ quarantine choices, and none of the pre-registered belief-by-barrier hypotheses was supported. Instead, randomized assignment to the high-barrier condition was associated with a lower quarantine rate, with closely agreeing estimates from the participant-level quasi-binomial and participant-day GEE analyses.

Three exploratory findings extend this central result. First, baseline perceived severity had a small positive, but non-confirmatory, association with quarantine behavior, similar in direction and magnitude to the earlier AUIB epigame [26]. Second, the estimated barrier response was larger among men than women, although the formal interaction was borderline and the two studies provide different forms of evidence for gender-related heterogeneity. Third, participants’ retrospective S3-Q3 assessments corresponded with their observed choices, especially in the high-barrier group. The simulated transmission trees were also strongly heterogeneous, while voluntary feedback identified both acceptable overall experience and specific technical and gameplay priorities. Together, these findings distinguish confirmed results within this trial from cross-study and post-outcome signals that warrant more focused prospective testing.

These findings distinguish two forms of validity that are not necessarily concordant. The S1–S2 associations provide evidence of indicator correspondence: participants carried relative differences in susceptibility, severity, self-efficacy, and perceived benefits from a real-life framing into the game. They do not, however, establish behavioral correspondence between virtual quarantine and protective behavior outside the game. Within the game, measured beliefs explained little of the variation in quarantine, whereas the experimentally assigned opportunity-cost condition produced a detectable behavioral difference. An epigame may therefore preserve the ordering of participants stated health beliefs while generating choices that are more responsive to the immediate decision environment than to those beliefs.

### 4.2 Belief Transfer Without Behavioral Transfer

The matched S1–S2 correlations and prospective ordinal models support construct-specific transfer rather than a general tendency to answer all belief questions similarly. Susceptibility showed the largest association, but all four domains remained associated after correction for multiple testing. The magnitudes were moderate rather than evidence of equivalence between real-life and in-game beliefs. Participants appeared to interpret the virtual pathogen through pre-existing belief structures, while also adapting those judgments to the game context. This is encouraging for the use of epigames to elicit psychologically meaningful attitudes, but it should not be interpreted as proof that the simulation reproduces real-world risk perception.

The absence of a corresponding relationship with quarantine behavior is substantively informative. The HBM describes several determinants that may contribute to protective action, but a stated belief need not translate directly into a repeated choice when the choice also carries immediate rewards, uncertain infection risk, social opportunities, and game-specific consequences. The weak severity trend was directionally consistent across an unadjusted correlation and the GEE model, but it did not survive multiplicity correction and was smaller under the quasi-binomial specification. It should therefore be treated as a hypothesis for future studies rather than evidence that severity independently drove quarantine in this trial.

The direction and magnitude of this signal are worth noting in the context of AUIB study. The participant-level Spearman association between baseline severity and quarantine rate was positive and small in both settings (AUIB: *ρ* = 0.132, unadjusted *p* = 0.0155; VinUni: *ρ* = 0.117, unadjusted *p* = 0.056). This agreement in direction does not convert these results into a confirmed effect, but it increases the value of specifying severity as a focused prospective hypothesis in the next implementation, using a reliable multi-item measure and a harmonized analysis across sites.

Measurement may also contribute to the disconnect. Each HBM domain was represented by a single six-point item, which limits reliability and may not capture the full construct or its moment-to-moment salience. Moreover, S2 measured beliefs early in the game, whereas quarantine was observed repeatedly as participants accumulated experience with rewards, infection, and other players. A stable survey response may consequently be a poor proxy for the changing considerations behind each daily decision. Future implementations could use validated multi-item scales and brief repeated belief measures timed before decisions, while preserving a prospectively specified analysis to avoid an excessive number of belief–behavior tests.

The exploratory S3-Q3 result provides a different and more limited form of behavioral correspondence. Participants who retrospectively described their game choices as more similar to intended real-life behavior also quarantined more often, particularly in G2. Because S3-Q3 was elicited after those choices, however, this pattern is compatible with participants reporting a self-description consistent with behavior they remembered, rather than a prior disposition producing that behavior. The group interaction is useful for hypothesis generation, but its mechanism and replicability require prospective testing.

### 4.3 Behavioral Response to the Barrier

The high-barrier group had 41% lower population-averaged odds of quarantine than the low barrier group, corresponding to an absolute difference of approximately 9.9 percentage points.

Agreement among the GEE, quasi-binomial, fractional-logit, and OLS sensitivity analyses indicates that this conclusion was not dependent on a single variance model or effect scale. The result supports the central experimental premise that an opportunity cost attached to remaining active can alter protective choices in a simulated epidemic.

The reward data adds an important qualification. The intended twofold contrast in points per contact was implemented closely, but participants had fewer qualifying contacts than anticipated. The realized difference in available contact rewards was therefore approximately 1.2 points per active day, substantially below the approximately 11-point tension used to motivate the design. The behavioral difference was nevertheless detectable. This pattern is compatible with participants responding to the salience or relative structure of the reward, but the trial did not independently randomize relative and absolute reward differences and cannot identify which feature produced the effect. Nor can the participant-level comparison of realized rewards be interpreted as a mediation analysis, because realized contact rewards depend on remaining active and on observed contacts after randomization. A future factorial experiment varying reward ratios and absolute point gaps separately would provide a direct test of these mechanisms.

### 4.4 Gender Pattern and Pre-Registered Hypotheses

Exploratory stratification suggested a larger barrier response among men: their quarantine rate was substantially lower in G2 than G1, whereas the estimated difference among women was smaller and imprecise. This pattern may be useful for designing future studies, but it does not establish that the barrier affected men and women differently. The formal gender-by-group interaction was borderline, the stratified comparison was exploratory, and the confidence interval for women included potentially meaningful effects. The appropriate interpretation is therefore that a gender-specific pattern was observed and requires prospective replication, not that women were unaffected or insensitive to cost.

The remaining pre-registered moderation hypotheses were consistently null under both final model families. In particular, the high barrier was not shown to act selectively among participants with uniformly low beliefs, belief-by-group interactions were jointly null, self-efficacy did not modify the barrier effect, and aggregate network degree did not predict or modify quarantine propensity. These null findings narrow the interpretation of the overall treatment effect. In this study, the barrier shifted average behavior, but the measured HBM constructs and network position did not identify who was most responsive to it.

### 4.5 Transmission Heterogeneity

The app-attributed transmission trees showed marked concentration: 71.5% of infected participants had no attributed secondary cases, 12.5% accounted for 80.0% of person-to-person transmissions, and the negative-binomial model estimated *k* = 0.176. The similarity of this dispersion estimate to the median reported for empirical COVID-19 transmission trees [29], together with the heterogeneous transmission described in another earlier epigame [20], shows that the platform can generate highly overdispersed transmission patterns through the interaction of simulated infection, observed proximity, and participant behavior. It does not establish that the game reproduced the biological or social mechanisms of COVID-19 superspreading. App attribution, finite-tree structure, end-of-study censoring, contact detection, interventions, and simulation parameters all shape the observed offspring distribution.

This result adds a network-level dimension to epigame validation. Belief correspondence evaluates whether stated constructs retain meaning across framings; the randomized contrast evaluates behavioral sensitivity to a designed incentive; and transmission-tree heterogeneity evaluates an emergent property of the coupled contact and simulation system. None is a substitute for the others. Future experiments should pre-specify transmission metrics, retain complete time-resolved attribution, and vary transmission and reward parameters separately to determine which design features generate concentration and whether behavioral interventions alter it.

### 4.6 Post-study Participant Feedback

Voluntary post-game feedback provided complementary implementation evidence (**Supplementary Table S8**; **Supplementary Figures S4–S5**). Among 215 submissions, the median overall experience rating was 4 of 5 (IQR 3–4), and the median ease-of-use rating was 8 of 10 (IQR 6–9). Points and rewards were the most frequently indexed enjoyed element (72/167 substantive submissions, 43.1%), whereas Bluetooth and contact detection were the most frequently indexed problem (73/175, 41.7%). Avatar and customization options led suggested improvements (57/150, 38.0%). This pattern identifies rewards and daily choices as salient enjoyed features, while contact detection and customization remain concrete development priorities. Because feedback was voluntary and themes were identified with overlapping bilingual keyword rules rather than formal qualitative coding, these results describe submitted feedback and should not be treated as representative prevalence estimates for all participants.

### 4.7 Strengths and Limitations[

This study combined randomized incentives, repeated observed decisions, prospectively measured beliefs, and passively measured contact networks in a socially embedded setting. The randomized design supports a causal interpretation of the assigned barrier effect within the analyzed trial population. The consistency of that estimate across aggregate and participant-day analyses is an additional strength. Treating S1–S2 correspondence prospectively and respecting the ordinal response scale also reduced reliance on exploratory model selection.

A number of limitations constrain our interpretation of the results. First, 283 of 357 randomized participants contributed a valid quarantine decision, and the available export could not distinguish withdrawal from failure to respond. Restriction to participants who engaged with the daily prompt may compromise the balance created by randomization if engagement was related jointly to treatment assignment and behavioral propensity. Second, participants contributed unequal numbers of decisions, ranging from 1 to 12. The quasi-binomial model retained these denominators, and the GEE modeled individual decisions, but neither approach eliminates selection caused by informative nonresponse. Third, repeated choices were strongly associated with participants, and substantial extra-binomial heterogeneity remained. Robust GEE and quasi-likelihood inference addressed uncertainty from these features, but the analyses do not identify the latent behavioral processes that generated them.

Fourth, Bluetooth proximity measurement and variable device performance may have introduced errors into contacts, rewards, and network degree. The null network findings could therefore reflect noise measurement or the limited ability of aggregate degree to represent the temporally varying opportunity cost faced at each decision. Fifth, the study was conducted on one university campus using virtual outcomes and lottery incentives. The results establish behavior within this experimental environment, not quarantine behavior during an actual epidemic. Sixth, S3 was completed by only 55.5% of the behavioral cohort, completion differed by randomized group, and completers contributed more daily decisions. S3 was also measured after gameplay and is vulnerable to recall, consistency, hypothetical, and post-randomization selection biases; its associations cannot establish prospective or causal effects. Finally, the gender-stratified analysis was exploratory, nonbinary and missing responses could not be included in the binary comparison, and subgroup estimates require replication in a study powered for interaction tests.

### 4.8 Implications and Conclusion

The principal implication is that validation of serious games should not be treated as a single question. A platform may demonstrate construct correspondence for stated beliefs while its observed choices remain governed by incentives and contingencies specific to the game. For behavioral research, survey realism and behavioral realism should therefore be evaluated separately. Experimental manipulation remains valuable precisely because they reveal how behavior changes under controlled conditions, but those responses should not automatically be interpreted as direct measures of real-world protective behavior.

The trends in the S3 survey reinforce this distinction. Retrospective correspondence between reported real-life intentions and observed game choices suggests that at least some participants recognized their behavior in the simulation, but it cannot establish prospective behavioral validity. A future protocol could measure intended responses before gameplay, collect brief repeated belief and intention measures before selected daily decisions, and reserve a post-game item for debriefing. This ordering would help distinguish prediction from recall-based consistency while reducing the ambiguity inherent in a single post-outcome self-assessment.

These findings provide a practical agenda for the next iteration. More reliable Bluetooth and contact detection would reduce measurement error in exposure, rewards, and transmission attribution; clearer notifications and decision windows could improve repeated response and attrition documentation; and richer avatar, shop, reward, and activity options could sustain engagement. Such refinements should not be assumed to make the observed severity or gender patterns statistically significant. Their scientific purpose is to improve treatment separation and measurement reliability so that prospectively specified tests can more decisively distinguish stable effects from chance or context-specific patterns.

Participants consistently transferred matched health beliefs into the simulated epidemic framing, yet those beliefs did not reliably predict quarantine choices. Randomized opportunity cost, by contrast, produced a robust average reduction in quarantine despite a smaller-than-planned realized reward difference. These results support epigames as tools for studying how stated beliefs and designed incentives coexist within an interactive epidemic environment. They also show why correspondence of attitudes alone is insufficient to establish behavioral validity. The study provides evidence of an evolving experimental platform: robust belief correspondence has now been observed across two settings (VinUni and AUIB), while severity, gender, retrospective self-assessment, and concentrated transmission define increasingly specific questions for confirmation. Future multisite trials should prospectively test these questions, manipulate absolute and relative reward magnitude separately, improve proximity sensing and engagement, measure beliefs closer to daily decisions, and harmonize documentation of participation and attrition. This progression supports epigames as iterative experimental tools, while keeping claims about real-world protective behavior contingent on direct prospective evidence.

## Data Availability

All data produced in the present study are available upon reasonable request to the authors

https://doi.org/10.5281/zenodo.23094742

## Ethics approval and consent

This study was approved by the UMass Chan Institutional Review Board (study ID 2465) and the VinUni-VinMec Institutional Ethical Review Board for Biomedical Research (approval no. 250/2025/CN/HDDD VMEC). All study procedures were carried out in accordance with Declaration of Helsinki. All participants were informed about the purpose of the study, assured of confidentiality, and provided digital consent prior to participation. Participation was voluntary, and respondents could withdraw at any time without consequence.

## Consent for publication

Not applicable.

## Availability of data and materials

Data available in Supplementary Materials and data sharing request to the corresponding author.

## Competing interests

The authors declare no competing interests.

## Funding

This study was supported partially by PD Fund AY25-26 (VG3862686/IO 30000132) awarded to PYW and CHS Trainee Research Grant AY25-26 (Trainee ID: V202301063, Program: BN, Grant ID: BN_CHS.TRP_2526_001) awarded to TLA supervised by PYW. The funding source played no role in the study’s design and conduct, data collection, management, analysis and interpretation, manuscript preparation, review, or approval, as well as the decision to submit the manuscript for publication.

## Author’s contributions

CA- conceptualization, methodology, data analysis, writing-original draft, writing-review & editing.

TLA- methodology, study design, data collection, writing-review & editing.

NLTM- methodology, study design, data collection, writing-review & editing.

HVNH- methodology, study design, data collection, data analysis, writing-review & editing.

ADNM- methodology, study design, data collection, writing-review & editing.

DLH- methodology, study design, data collection, writing-review & editing.

MDH- methodology, study design, data collection, writing-review & editing.

DK- initial conceptualization.

JPG – conceptualization, writing-review & editing.

PYW- conceptualization, methodology, supervision, writing-original draft, writing-review & editing.

All authors contributed to the final version of the manuscript and approved submission. All authors had full access to all the data in the study, and the corresponding author had final responsibility to submit for publication.

## Acknowledgements

We thank all VinUni participants for their time and contribution to this study. We are also grateful to the student investigators TLA, NLTM, HVNH, ADNM, DLH, MDH for their assistance with study preparation, participant recruitment and data collection under the supervision of PYW and CA. Special thanks to DK for connecting and introducing Epigame trial in VinUni.

## Notes

### Competing Interest Statement

The authors have declared no competing interest.

### Author Declarations

The IRB of UMass Chan Medical School and VinUni-VinMec gave ethical approval for this work

### Summary of Updates

Corrected the names of some of the co-authors, and made some minor typo edits.

