## Supplementary materials for "Evaluating Behavioral Parallelism and Cost Incentives in Voluntary Quarantine: A Randomized App-Based Epidemic Game at VinUniversity in Hanoi, Vietnam" (from another version of this preprint: version 1, 2026-10-08)

### Supplementary Methods

### Exploratory Exit-Survey Analysis

The exit survey (S3) was administered after gameplay. The four substantive items used six-point response scales:

- **S3-Q1, perceived realism:** "How well do you think the game represents infectious disease transmission in real life?" Higher values indicate greater perceived realism.
- **S3-Q2, concern:** "Were you worried about getting virtually infected during the game?" Higher values indicate greater concern.
- **S3-Q3, retrospective real-life correspondence:** "How closely did your quarantine decisions during the game reflect what you would do in a real-life outbreak?" Higher values indicate greater reported correspondence.
- **S3-Q4, difficulty understanding information:** "How difficult was it to understand the information you received during the game?" Higher values indicate greater difficulty, not better understanding.

All four items were available for the same 157 participants, who contributed 1,238 valid participant-days. Participant-level Spearman correlations described unadjusted monotonic associations with quarantine rate. Group-adjusted associations were estimated separately for each item using (1) participant-day binomial generalized estimating equations (GEE) with participant clustering, exchangeable working correlation, and robust standard errors and (2) participant-level binomial generalized linear models of quarantine proportion, weighted by each participant's number of valid decisions and fitted with Pearson scaling (scale='X2'). Separate interaction models included the item, randomized group, and their interaction. Items were mean-centered within the S3 sample; odds ratios (ORs) represent a one-point increase on the original scale.

Holm correction was applied separately across the four items for each of five declared families: Spearman associations, group-adjusted GEE associations, group-adjusted quasi-binomial associations, GEE item-by-group interactions, and quasi-binomial item-by-group interactions. Arm-specific slopes decompose the interaction models and were not treated as additional discovery families. These analyses were not pre-registered and are post-outcome: they assess retrospective correspondence, not prospective prediction or causation.

### Supplementary Results

### S3 Completion and Analysis Sample

Complete S3 responses were available for 82 of 132 G1 participants (62.1%) and 75 of 151 G2 participants (49.7%). Completion differed by randomized group ($\chi_1^2 = 4.42, p = 0.0355$). Completers contributed more valid daily decisions than non-completers (means 7.89 versus 4.64; Mann–Whitney $p < 0.001$). Results may therefore be affected by differential engagement and post-randomization selection.

### Supplementary Table S1. Exploratory Associations Between S3 Responses and Quarantine Behavior

#### Panel A. Participant-level Spearman correlations

| Item | Construct | *N* | *ρ* | Raw *p* | Holm *p* |
|---|---|---|---|---|---|
| S3-Q1 | Perceived realism | 157 | 0.148 | 0.0648 | 0.1295 |
| S3-Q2 | Concern about virtual infection | 157 | 0.162 | 0.0427 | 0.1282 |
| S3-Q3 | Retrospective real-life correspondence | 157 | 0.283 | 0.000337 | 0.00135 |
| S3-Q4 | Difficulty understanding information | 157 | 0.074 | 0.3603 | 0.3603 |

#### Panel B. Group-adjusted and group-stratified regression estimates

| Item | Model | Group-adjusted OR (95% CI) | Raw / Holm *p* | G1 OR (95% CI); *p* | G2 OR (95% CI); *p* | Interaction ratio of ORs (95% CI) | Raw / Holm interaction *p* |
|---|---|---|---|---|---|---|---|
| S3-Q1 | GEE | 1.26 (1.04–1.53) | 0.0179 / 0.0538 | 1.29 (0.99–1.68); 0.0602 | 1.23 (0.93–1.62); 0.1515 | 0.95 (0.65–1.40) | 0.7992 / 1.0000 |
| S3-Q1 | Quasi-binomial | 1.31 (1.09–1.58) | 0.00486 / 0.0146 | 1.34 (1.05–1.71); 0.0176 | 1.26 (0.94–1.70); 0.1276 | 0.94 (0.64–1.38) | 0.7627 / 1.0000 |
| S3-Q2 | GEE | 1.15 (0.96–1.39) | 0.1378 / 0.2757 | 1.04 (0.84–1.29); 0.7312 | 1.36 (0.97–1.92); 0.0759 | 1.31 (0.88–1.96) | 0.1882 / 0.5647 |
| S3-Q2 | Quasi-binomial | 1.19 (1.01–1.41) | 0.0366 / 0.0733 | 1.10 (0.89–1.36); 0.3564 | 1.37 (1.03–1.82); 0.0325 | 1.24 (0.87–1.76) | 0.2413 / 0.7239 |
| S3-Q3 | GEE | 1.41 (1.16–1.71) | 0.000480 / 0.00192 | 1.13 (0.88–1.45); 0.3338 | 1.88 (1.41–2.50); 0.000015 | 1.66 (1.14–2.42) | 0.00845 / 0.0338 |
| S3-Q3 | Quasi-binomial | 1.39 (1.16–1.67) | 0.000321 / 0.00128 | 1.13 (0.90–1.43); 0.2831 | 1.83 (1.37–2.44); 0.000046 | 1.61 (1.11–2.33) | 0.0117 / 0.0469 |
| S3-Q4 | GEE | 1.13 (0.95–1.33) | 0.1680 / 0.2757 | 1.09 (0.87–1.38); 0.4478 | 1.17 (0.93–1.49); 0.1859 | 1.07 (0.77–1.50) | 0.6741 / 1.0000 |
| S3-Q4 | Quasi-binomial | 1.07 (0.89–1.29) | 0.4485 / 0.4485 | 1.05 (0.83–1.33); 0.6796 | 1.11 (0.82–1.51); 0.4875 | 1.06 (0.72–1.56) | 0.7707 / 1.0000 |

***Notes:*** All ORs represent a one-point increase in the corresponding S3 response. Group-adjusted estimates come from item-plus-group models. G1 and G2 slopes and the interaction ratio come from item-by-group models; the interaction is the G2 slope divided by the G1 slope. GEE models include 1,238 participant-days from 157 participants. Quasi-binomial interaction-model Pearson scales were 3.83 (Q1), 4.04 (Q2), 3.55 (Q3), and 3.90 (Q4). Raw and Holm-adjusted *p*-values are shown because the analyses are exploratory. The apparent Q1 association was statistically significant only in the quasi-binomial model after Holm correction; Q3 was the only item consistently associated across descriptive, GEE, and quasi-binomial analyses and the only item with a supported interaction in both model families.

#### Final Models for the Pre-Registered Hypotheses

The final hypothesis analyses used two complementary specifications. Participant-day GEE models used a binomial family, logit link, participant clustering, exchangeable working correlation, and robust standard errors. Participant-level quasi-binomial models used quarantine rate as the outcome, the number of valid decisions as the binomial variance weight, and Pearson scaling. S1 and S2 belief items were mean-centered. H1 and H1b were evaluated with linear contrasts for the protocol-defined profile scoring 1 on all four belief items, standardized over the analysis sample's gender distribution. H2 and H2b were four-degree-of-freedom joint tests of the belief-by-group interactions. G1 (low barrier) was the reference group.

The overall randomized arm effect was distinct from H1/H1b. In the full cohort, G2 had lower odds of quarantine than G1 in both the GEE (OR 0.59, 95% CI 0.40–0.86, $p = 0.0059$) and quasi-binomial model (OR 0.59, 95% CI 0.40–0.86, $p = 0.0055$; 283 participants and 1,823 decisions). This supports an average barrier effect, but it does not support the proposed belief-dependent mechanisms.

### Supplementary Table S2. Final Tests of Pre-Registered Hypotheses

| Hypothesis | Analysis sample | GEE result | Quasi-binomial result | Decision |
|---|---|---|---|---|
| H1: barrier effect for S1 skeptic profile | 251 participants; 1,642 decisions | OR 0.50 (0.11–2.34), $z = -0.88$, $p = 0.378$ | OR 0.57 (0.10–3.07), $z = -0.66$, $p = 0.509$ | Not supported |
| H1b: barrier effect for S2 skeptic profile | 235 participants; 1,569 decisions | OR 1.72 (0.30–9.92), $z = 0.61$, $p = 0.545$ | OR 1.62 (0.27–9.65), $z = 0.53$, $p = 0.599$ | Not supported; point estimate opposed prediction |
| H2: joint S1 belief-by-group moderation | 251 participants; 1,642 decisions | Wald $\chi^2_4 = 1.10$, $p = 0.895$ | Wald $\chi^2_4 = 1.50$, $p = 0.826$ | Not supported |
| H2b: joint S2 belief-by-group moderation | 235 participants; 1,569 decisions | Wald $\chi^2_4 = 2.82$, $p = 0.589$ | Wald $\chi^2_4 = 4.43$, $p = 0.351$ | Not supported |
| H3: S1 self-efficacy-by-group interaction | 251 participants; 1,642 decisions | OR 0.84 (0.57–1.24), $z = -0.87$, $p = 0.382$ | OR 0.85 (0.61–1.18), $z = -0.98$, $p = 0.329$ | Not supported |
| H3b: S2 self-efficacy-by-group interaction | 235 participants; 1,569 decisions | OR 0.80 (0.53–1.21), $z = -1.06$, $p = 0.292$ | OR 0.87 (0.57–1.32), $z = -0.67$, $p = 0.505$ | Not supported |
| H4: node-degree-by-group interaction | 283 participants; 1,823 decisions | $\beta = 0.0052$, OR 1.005 (approximately 1.00–1.02), $z = 0.96$, $p = 0.338$ | $\beta = 0.0055$, OR 1.006 (approximately 1.00–1.02), $z = 1.01$, $p = 0.315$ | Not supported; direction opposed prediction |
| H5: women versus men, S1-adjusted | 251 participants; 1,642 decisions | OR 1.25 (0.82–1.89), $z = 1.05$, $p = 0.295$ | OR 1.18 (0.78–1.77), $z = 0.78$, $p = 0.437$ | Not supported |
| H5: women versus men, S2-adjusted | 235 participants; 1,569 decisions | OR 1.22 (0.80–1.85), $z = 0.93$, $p = 0.353$ | OR 1.19 (0.79–1.79), $z = 0.82$, $p = 0.412$ | Not supported |

***Notes:*** ORs below 1 were predicted for the barrier contrasts and the H3/H3b and H4 interaction terms. The H5 rows test the gender main effect within the corresponding multivariable belief model. H5 does not test the exploratory gender-by-group interaction reported in the main text. The H1/H1b confidence intervals are linear-contrast intervals on the log-odds scale transformed to ORs.

### Supplementary Table S3. Belief-by-Group Coefficients Underlying H2 and H2b

| Survey and interaction | GEE $\beta$ | GEE OR (95% CI) | GEE $p$ | Quasi-binomial $\beta$ | Quasi-binomial OR (95% CI) | Quasi-binomial $p$ |
|---|---|---|---|---|---|---|
| S1 susceptibility × G2 | 0.026 | 1.03 (0.75–1.41) | 0.872 | 0.053 | 1.05 (0.74–1.51) | 0.773 |
| S1 severity × G2 | -0.058 | 0.94 (0.66–1.34) | 0.747 | -0.141 | 0.87 (0.59–1.29) | 0.482 |
| S1 self-efficacy × G2 | -0.172 | 0.84 (0.57–1.24) | 0.382 | -0.164 | 0.85 (0.61–1.18) | 0.329 |
| S1 perceived benefits × G2 | 0.193 | 1.21 (0.77–1.92) | 0.409 | 0.210 | 1.23 (0.78–1.96) | 0.375 |
| S2 susceptibility × G2 | -0.215 | 0.81 (0.55–1.19) | 0.283 | -0.313 | 0.73 (0.50–1.07) | 0.109 |
| S2 severity × G2 | -0.136 | 0.87 (0.60–1.28) | 0.488 | -0.122 | 0.89 (0.63–1.25) | 0.490 |
| S2 self-efficacy × G2 | -0.220 | 0.80 (0.53–1.21) | 0.292 | -0.144 | 0.87 (0.57–1.32) | 0.505 |
| S2 perceived benefits × G2 | 0.101 | 1.11 (0.72–1.70) | 0.645 | 0.101 | 1.11 (0.73–1.67) | 0.631 |

***Notes:*** Each row is the difference in the belief slope between G2 and G1. The four coefficients within S1 and S2 were tested jointly for H2 and H2b, respectively; those joint tests are reported in Supplementary Table S2. Individual coefficients were not interpreted as separate confirmations of the joint hypotheses.

### Variance and Within-Participant Dependence Diagnostics

The original Poisson count and ordinary weighted-binomial specifications treated repeated decisions as conditionally independent. Their Pearson statistics were substantially larger than their residual degrees of freedom, implying standard errors approximately 1.7–1.9 times those from an equidispersed model (Supplementary Table S4). Final quasi-binomial scales remained close to 3 across model families, while exchangeable GEE working correlations ranged from 0.37 to 0.42.

### Supplementary Table S4. Overdispersion in the Original Model Specifications

| Original model | Participants | Pearson $\chi^2/df$ | Implied SE multiplier |
|---|---|---|---|
| Poisson arm-only quarantine-count model | 283 | 3.32 | 1.82 |
| Poisson S1 belief-and-group interaction model | 251 | 3.53 | 1.88 |
| Poisson S2 belief-and-group interaction model | 235 | 3.26 | 1.81 |
| Poisson node-degree-and-group interaction model | 283 | 3.42 | 1.85 |

| Original model | Participants | Pearson $\chi^2/df$ | Implied SE multiplier |
|---|---|---|---|
| Weighted-binomial S2 HBM model | 235 | 3.10 | 1.76 |
| Weighted-binomial gender-by-group model | 251 | 3.02 | 1.74 |

***Notes:*** The SE multiplier is $\sqrt{\chi^2/df}$ and approximates how much the ordinary model understated uncertainty when the mean model is retained. Final quasi-binomial Pearson scales were 3.15 for the S1 interaction model, 3.07 for the S2 interaction model, 3.05 for the node-degree interaction model, 3.08 for the additive S1 HBM model, and 3.10 for the additive S2 HBM model. Corresponding GEE working correlations were 0.419, 0.369, 0.399, 0.409, and 0.392.

Participant-level rates also showed more boundary mass and variability than expected under a binomial model that retained each participant's observed number of prompts (Supplementary Figure S1). In addition, consecutive choices showed strong serial dependence (Supplementary Table S5).

### Supplementary Table S5. Boundary Mass and Consecutive-Decision Dependence

| Diagnostic | Observed result | Independent binomial reference |
|---|---|---|
| Participants with quarantine rate 0 | 126/283 (44.5%) | 23.1% |
| Participants with quarantine rate 1 | 20/283 (7.1%) | 2.7% |
| SD of participant quarantine rate | 0.314 | 0.217 |
| Observed/null variance ratio | 2.09 | 1.00 |
| Consecutive-day decision pairs | 1,067 | – |
| $P(Q_t = 1 \mid Q_{t-1} = 0)$ | 0.141 | – |
| $P(Q_t = 1 \mid Q_{t-1} = 1)$ | 0.626 | – |
| Lag-1 odds ratio | 10.23 | 1.00 |
| Transition-table test | $\chi^2_1 = 242.4, p = 1.15 \times 10^{-54}$ | – |

***Notes:*** The binomial reference was generated separately for each participant using that participant's observed number of prompts and the pooled quarantine probability. Consecutive pairs required decisions on adjacent study days. Stable participant heterogeneity and within-participant serial dependence are not cleanly separable in these descriptive diagnostics; both invalidate ordinary independent-trial standard errors and motivate the final quasi-binomial and GEE analyses.

### Transmission-Tree Heterogeneity

Exploratory superspreader analyses used the complete app-attributed transmission graph rather than the cleaned daily infection series. This preserved all available infector–infectee chains and yielded three rooted trees containing 288 unique infected participants and 285 attributed person-to-person transmissions. The offspring count was each node's out-degree. Intercept-only Poisson and negative-binomial models were fitted to this distribution; the negative-binomial dispersion parameter was calculated as $k = 1/\alpha$. A superspreader was defined as a case above the 99th percentile of a Poisson distribution with the observed mean, which gave a threshold of more than four secondary cases. These analyses were exploratory.

### Supplementary Table S6. App-Attributed Transmission-Tree Heterogeneity

| Diagnostic | Result |
|---|---|
| Transmission trees | 3 |
| Infected participants | 288 |

| Diagnostic | Result |
|---|---|
| Attributed person-to-person transmissions | 285 |
| Mean secondary cases | 0.990 |
| Variance of secondary cases | 6.538 |
| No attributed secondary cases | 206/288 (71.5%) |
| Maximum secondary cases | 20 |
| Negative-binomial dispersion, $k$ | 0.176 |
| Negative-binomial AIC | 680.29 |
| Poisson AIC | 1189.77 |
| Superspreaders, $>$ 4 secondary cases | 19/288 (6.6%) |
| Transmissions attributed to superspreaders | 171/285 (60.0%) |
| Minimum group accounting for at least 80% of transmission | 36/288 (12.5%) |
| Transmissions attributed to that group | 228/285 (80.0%) |

***Notes:*** The mean secondary-case count is $(n-1)/n$ within each completed rooted tree and is therefore constrained toward 1; it is not an estimate of biological $R_0$. The substantially lower negative-binomial AIC describes the observed offspring distribution better than the equidispersed Poisson model. The estimated $k = 0.176$ is close to the median $k = 0.14$ reported for COVID-19 trees in OutbreakTrees, but direct equivalence is not implied. App attribution, finite-tree and end-of-study censoring, behavioral interventions, network structure, and simulation parameters all affect these estimates. The tree denominator differs from the 191 daily infection events in the main analysis because complete chains were retained before cleaned behavioral exclusions.

### Effect of point score

The barrier was manipulated through the per-contact bonus (1 pt/contact in G1, 2 pts/contact in G2, contacts >= 5 min, capped at 15/day). Here we verify that the reward schedule the participants experienced matched the design and quantify the realized economic barrier, which is the points forgone by choosing to quarantine for a day.

### Supplementary Table S7. Realized reward differential

| Item | G1 (low barrier) | G2 (high barrier) | Ratio | P-value |
|---|---|---|---|---|
| Score | 46.704 | 47.572 | 1.02 | 0.414 |
| Scoring contacts | 5 | 4 | 0.8 | 0.72 |
| Contact points | 3.886 | 7.368 | 1.9 | 0.0817 |
| Points per contact | 0.959 | 1.973 | 2.06 | 6.40E-29 |
| Scoring contacts per active day | 1.458 | 1.333 | 0.91 | 0.447 |
| Realized barrier | 1.141 | 2.325 | 2.04 | 0.0787 |

### Voluntary Post-Game Feedback

The separate post-game feedback form received 215 voluntary submissions: 80 through the English-language pathway and 135 through the Vietnamese-language pathway. The form was not linked to the behavioral analysis for inference. Names, email addresses, application identifiers, timestamps, and verbatim comments were excluded from all analytic exports.

Closed-ended items rated overall experience on a five-point scale and ease of application use on a ten-point scale. Favorable ratings were defined descriptively as at least 4 of 5 and at least 8 of 10, respectively. English and Vietnamese free-text fields were paired by prompt and normalized for reproducible bilingual keyword matching. Related prompts were combined into three domains: enjoyed elements, reported problems, and suggested improvements. A submission could match multiple themes but was counted at most once per theme within each domain. Percentages use submissions with at least one substantive comment in the corresponding domain as their denominator. These overlapping themes are descriptive indexes of mentions, not sentiment scores or validated qualitative codes.

**Supplementary Table S8. Ratings and Leading Themes in Voluntary Post-Game Feedback**

| Domain | Rating or theme | Result |
|---|---|---:|
| Overall experience | Five-point rating | Median 4 (IQR 3–4); 146/215 (67.9%) rated 4 or 5 |
| Ease of application use | Ten-point rating | Median 8 (IQR 6–9); 115/215 (53.5%) rated 8–10 |
| Enjoyed elements | Points and rewards | 72/167 (43.1%) |
| Enjoyed elements | Quarantine and daily choice | 62/167 (37.1%) |
| Enjoyed elements | Social contact and Bluetooth | 38/167 (22.8%) |
| Reported problems | Bluetooth and contact detection | 73/175 (41.7%) |
| Reported problems | Shop or items unavailable | 50/175 (28.6%) |
| Reported problems | Notifications and decision window | 44/175 (25.1%) |
| Suggested improvements | Avatar and customization | 57/150 (38.0%) |
| Suggested improvements | Rewards, shop, and items | 47/150 (31.3%) |
| Suggested improvements | More challenges and activities | 35/150 (23.3%) |

*Notes:* Theme denominators are the numbers of submissions containing at least one substantive response within the corresponding domain. Percentages therefore quantify mentions among substantive submissions, not among all study participants. Theme matches could overlap. Keyword rules included English and Vietnamese terms and did not assign uncoded text to themes; the results are consequently non-exhaustive. No response-level or identifying data are reported.

### Supplementary Figures

#### Supplementary Figure S1. Within-Participant Heterogeneity and Serial Dependence

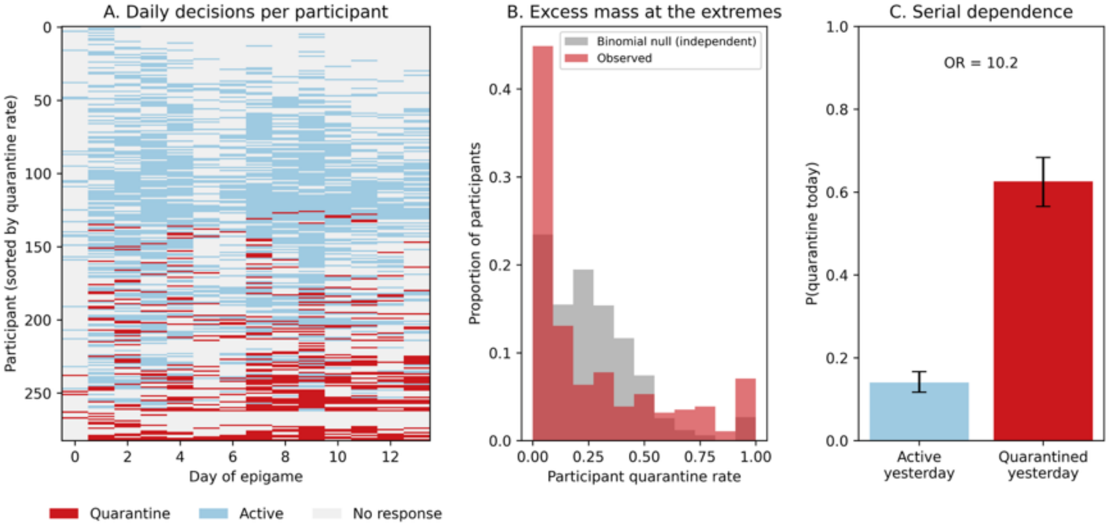


*Supplementary Figure S1. Within-participant heterogeneity and serial dependence.*

**Supplementary Figure S1.** Repeated quarantine decisions and departures from an independent-binomial model. (A) Participant-by-day decision raster, with participants sorted by observed quarantine rate. (B) Distribution of observed participant quarantine rates compared with a simulated binomial reference that retains each participant's number of prompts, showing excess mass at rates 0 and 1. (C) Probability of quarantine following an active versus quarantine decision on the preceding study day; error bars are exact binomial 95% confidence intervals. The lag-1 odds ratio was 10.23. Panels B and C represent stable between-participant heterogeneity and within-participant momentum, respectively; they are descriptive and are not assumed to identify separate causal processes.

#### Supplementary Figure S2. App-Attributed Transmission Trees

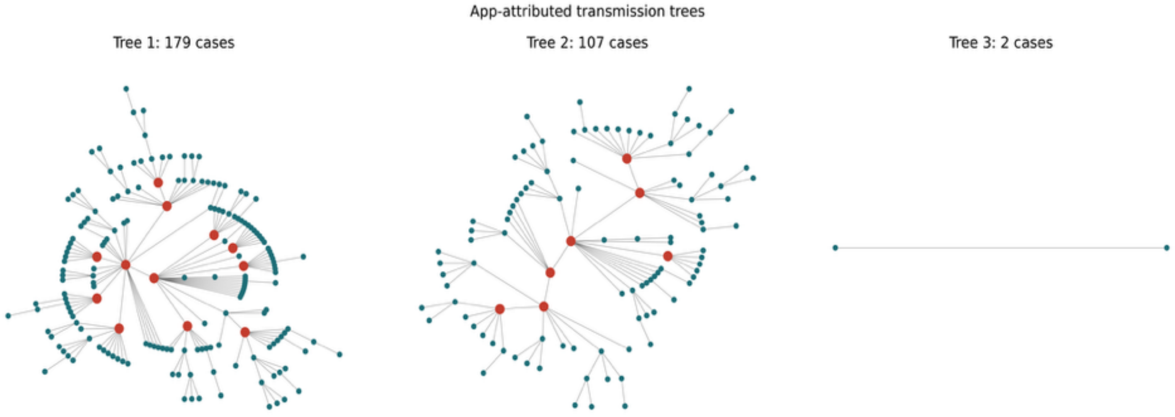


*Supplementary Figure S2. App-attributed transmission trees.*

**Supplementary Figure S2.** The three connected components of the complete app-attributed transmission graph, ordered by size. Nodes represent infected participants and edges represent attributed person-to-person transmission. Red nodes generated more than four secondary cases and met the superspreader definition; teal nodes did not. The trees contained 179, 107, and 2 cases. Layouts display topology rather than infection timing or physical distance.

### Supplementary Figure S3. Offspring Heterogeneity and Concentration of Transmission

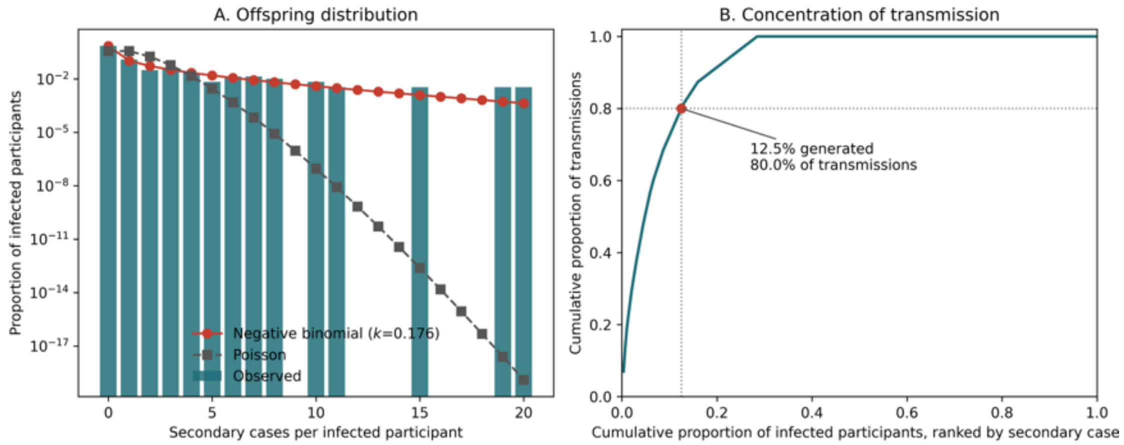


*Supplementary Figure S3. Offspring heterogeneity and concentration of transmission.*

**Supplementary Figure S3.** Heterogeneity in app-attributed secondary transmission across all three trees. (A) Observed offspring distribution compared with fitted negative-binomial ($k = 0.176$) and Poisson distributions; the vertical axis is logarithmic. (B) Cumulative share of the 285 attributed person-to-person transmissions after ranking all 288 infected participants by their number of secondary cases. The smallest group reaching 80% comprised 36 participants (12.5%). These are descriptive properties of a finite simulated outbreak and are not estimates of biological transmissibility.

### Supplementary Figure S4. Voluntary Post-Game Ratings

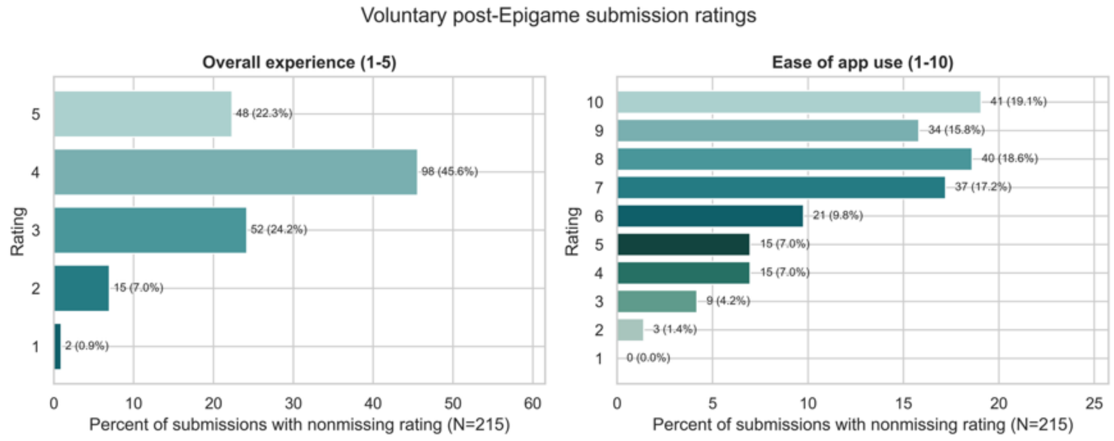


*Supplementary Figure S4. Voluntary post-game ratings.*

**Supplementary Figure S4.** Distribution of ratings across 215 voluntary post-game feedback submissions. (A) Overall experience on a five-point scale. (B) Ease of application use on a ten-point scale. Bars retain every response category; percentages use all 215 submissions as the denominator.

### Supplementary Figure S5. Themes in Voluntary Post-Game Feedback

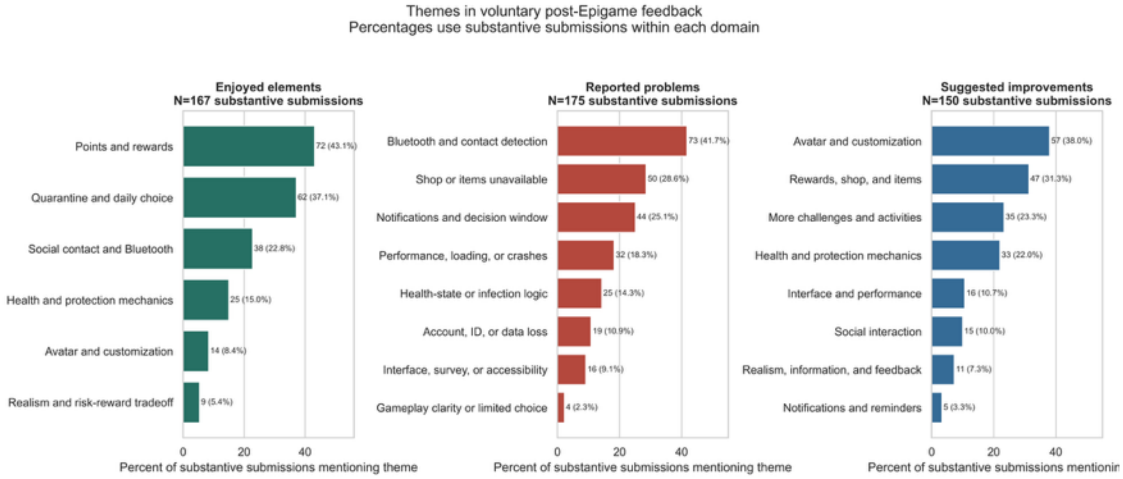


*Supplementary Figure S5. Themes in voluntary post-game feedback.*

**Supplementary Figure S5.** Reproducibly indexed bilingual free-text themes for enjoyed elements, reported problems, and suggested improvements. Percentages use submissions with at least one substantive comment in the displayed domain as the denominator ($N = 167$, 175, and 150, respectively). A submission could match more than one theme. The keyword-based themes are descriptive and non-exhaustive and do not measure sentiment intensity or qualitative importance.
